# Climate and land-use shape the spread of zoonotic yellow fever virus

**DOI:** 10.1101/2022.08.25.22278983

**Authors:** Sarah C. Hill, Simon Dellicour, Ingra M. Claro, Patricia C. Sequeira, Talita Adelino, Julien Thézé, Chieh-Hsi Wu, Filipe Romero Rebello Moreira, Marta Giovanetti, Sabrina L. Li, Jaqueline G. de Jesus, Felipe J. Colón-González, Heather R. Chamberlain, Oliver Pannell, Natalia Tejedor-Garavito, Fernanda de Bruycker-Nogueira, Allison A. Fabri, Maria Angélica Mares-Guia, Joilson Xavier, Alexander E. Zarebski, Arran Hamlet, Maria Anice Mureb Sallum, Antonio C. da Costa, Erika R. Manuli, Anna S. Levin, Luís Filipe Mucci, Rosa Maria Tubaki, Regiane Maria Tironi de Menezes, Juliana Telles de Deus, Roberta Spinola, Leila Saad, Esper G. Kallas, G.R. William Wint, Pedro S. Peixoto, Andreza Aruska de Souza Santos, Jane P. Messina, Oliver J. Brady, Andrew J. Tatem, Marc A. Suchard, Jairo A. Mendez-Rico, André Abreu, Renato Santana Aguiar, Oliver G. Pybus, Guy Baele, Philippe Lemey, Felipe Iani, Mariana S. Cunha, Ana M. Bispo de Filippis, Ester C. Sabino, Nuno R. Faria

## Abstract

Zoonotic viruses that originate in wildlife harm global human health and economic prosperity^1^. Understanding virus transmission at the human-animal-environment interface is a key component of pandemic risk-reduction^2,3^. Zoonotic disease emergence is highest in biodiverse, tropical forests undergoing intensive land-use change^4,5^. Phylodynamic analyses of virus genomes can powerfully test epidemiological hypotheses, but are rarely applied to viruses of animals inhabiting these habitats. Brazil’s densely-populated Atlantic Forest and Cerrado region experienced in 2016–2021 an explosive human outbreak of sylvatic yellow fever, caused by repeated virus spillover from wild neotropical primates^6^. Here we use yellow fever virus (YFV) genome sequences and epidemiological data from neotropical primates, humans, and mosquito vectors to identify the environmental, demographic, and climatic factors determining zoonotic virus spread. Using portable sequencing approaches we generated 498 YFV genomes, resulting in a well-sampled dataset of zoonotic virus genomes sampled from wild mammals. YFV dispersal velocity was slower at higher elevation, in colder regions, and further away from main roads. Virus lineage dispersal was more frequent through wetter areas, areas with high neotropical primate density and through landscapes covered by mosaic vegetation. Higher temperatures were associated with higher virus effective population sizes, and peaks of transmission in warmer, wetter seasons were associated with higher virus evolutionary rates. Our study demonstrates how zoonotic disease transmission is linked to land-use and climate, underscoring the need for One-Health approaches to reducing the rate of zoonotic spillover.

## Main

Wildlife are a major source of zoonotic emerging infectious diseases^1^. The Quadripartite Commitment undertaken by the World Health Organization (WHO), World Organisation for Animal Health (WOAH), Food and Agriculture Organization of the United Nations (FAO) and United Nations Environment Programme (UNEP) advocates the need for stronger early-warning systems of zoonotic disease emergence^7^. An important pre-requisite for targeting activities that reduce spillover, including pre-emptive vaccination, is knowing how viruses spread within wildlife reservoirs. Yet for most zoonotic viruses we have limited understanding of what causes epizootic dynamics, such as the duration of persistence within a wildlife host population or geographic routes of virus movement.

Many RNA viruses evolve rapidly enough to accumulate genomic diversity that contains information about transmission processes during an outbreak^8,9^. Phylodynamic analyses of virus genomes support the identification— and measurement of the importance of— variables correlated with changes in outbreak size, dispersal velocity, and regional source-sink dynamics^10–13^. Such approaches have been applied extensively to human-to-human transmission of epidemics of zoonotic origin (*e.g*., SARS-CoV-2^14^ and Ebola virus^11^). However, this attention to “post-spillover” human-focused transmission reveals little about the processes underlying epidemic origins and transmission in the animal reservoir. We previously lacked sufficient zoonotic virus genomes from wild mammalian host reservoirs to fill this gap.

Yellow fever virus (YFV) is a zoonotic, single-stranded RNA virus belonging to the genus *Flavivirus*^15^. In the Americas, YFV is maintained primarily within a sylvatic transmission cycle by mosquitos from the *Haemagogus* and *Sabethes* genera, which breed in tree cavities and feed on a range of vertebrates^16–20^. These mosquitos typically transmit YFV among neotropical primates (NPs), but can also bite and infect humans in the vicinity of ongoing epizootics^21,22^. Brazil is currently recovering from its largest outbreak of YFV in >75 years. The outbreak’s high human morbidity resulted from virus re-emergence in coastal areas of southeast Brazil, where YFV was previously absent and where vaccination was not routinely recommended^23^. Over 2000 human cases of yellow fever were confirmed related to extensive epizootic transmission in the South, Southeast and Northeast regions of Brazil between 2016 and 2021^24^. Dramatic declines in NPs were observed concurrently during the outbreak, including for critically endangered and vulnerable species^25^. The worst affected region contains three of South America’s most populous cities (São Paulo, Rio de Janeiro, and Belo Horizonte) alongside the Atlantic Forest, a heavily deforested and fragmented biome that nevertheless hosts one of the highest diversity of vertebrates worldwide^26,27^. The outbreak therefore represents an opportunity to test hypotheses about factors that drive zoonotic virus spread in an extensive animal-human interface^5^.

Previous studies have used YFV genomes to spatially reconstruct YFV spread^6,28–30^ but have not directly tested hypotheses regarding the key environmental drivers of transmission and persistence. Here, we quantify the association of demographic, climatic and environmental factors with heightened YFV transmission in Brazil and investigate factors supporting inter-epidemic persistence. We apply a suite of phylodynamic and phylogeographic models to a comprehensive and representative dataset of 705 YFV genomes from 2016–2019, which represent nearly 15% of confirmed infected humans or NPs. This genomic sampling fraction is comparable to that of the best-sampled human outbreaks, including Ebola virus^11^ in West Africa (5%) and SARS-CoV-2 in the UK^31^ (15%), and is therefore an exceptional genetic dataset for a zoonotic outbreak within a wild mammalian host. Our findings reveal the factors that must be considered in efforts to reduce morbidity in future YFV epidemics, and global amplification of zoonotic diseases^32–36^.

### Overview of YFV spread in Brazil, 2016–2019

We used portable MinION sequencing technology to generate 498 complete or near-complete (>70% of coding region) genomes from the outbreak in Brazil, tripling the number of genomes available from this event^28^. A total of 71% of the newly generated genomes were sampled from NPs and mosquitos (**Figure 1**).

**Figure 1:**
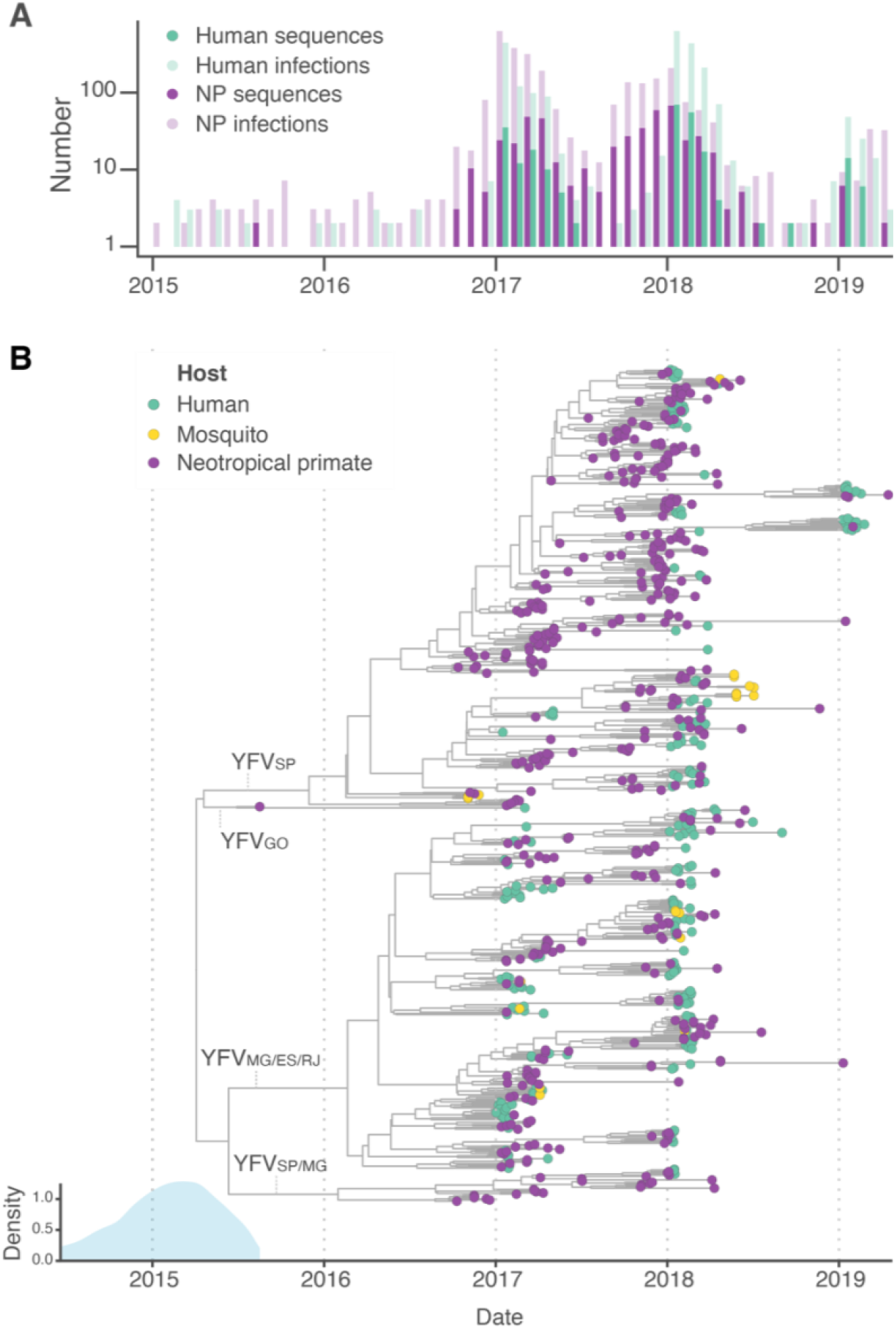
**A:** Sampling of confirmed YFV infections. Pale colours indicate unsequenced YFV confirmed infections per month in NPs (purple) and humans (green), and darker colours indicate sequenced infections per month and host (i.e., the total height of each bar represents the number of confirmed infections per month and host). **B:** Maximum clade credibility (MCC) tree of Brazilian YFV outbreak clade obtained during continuous phylogeographic analysis. Colours at tips denote host type. Two major and two minor clades are labelled by main federal states of circulation; see **Extended Data Figure 1A** for tree with all tips coloured by federal state. Density plot on bottom left shows marginal density of estimated TMRCA of the root, with the lower tail truncated to 95% HPD region (minimum value; 2013.2).

When combined with curated publicly-available data, all but one of the genomes sampled in Brazil since 2010 fell in a single monophyletic clade within the overall diversity of the South American I YFV genotype (maximum likelihood phylogeny bootstrap score = 100, **Extended Data Figure 1**); we focused on this clade in subsequent analyses. The previously reported outlier sequence^37^ was sampled in 2017 in northwestern Goiás state and is geographically and genetically distinct from the other sequenced samples considered here. The time of the most recent common ancestor (TMRCA) of the monophyletic YFV sequences from southeast and bordering areas of Brazil (federal states of São Paulo (SP), Minas Gerais (MG), Espírito Santo (ES), Rio de Janeiro (RJ), Bahia (BA) and Goiás (GO)) is January 2015 (95% highest posterior density interval [HPD], June 2014 to August 2015). This estimate is slightly earlier than from previous studies^3 16,30^ because we include additional sequences from more geographic locations and therefore capture earlier transmission events.

Following emergence, one major clade circulated in humans and NPs in São Paulo state (SP) and nearby municipalities in Minas Gerais (MG) whilst a second clade spread primarily in MG, Espírito Santo (ES), and Rio de Janeiro (RJ) (respectively denoted YFVSP and YFVMG/ES/RJ on **Figure 1**, and visualised spatially in **Figure 2)**. Two smaller clades exist that represent (i) transmission focused in Goiás (GO), and (ii) a second introduction to MG and SP from which we sampled fewer YFV infections (respectively marked as YFVGO and YFVSP/MG on **Figure 1B** and coloured by state on **Extended Data Figure 1A**). Virus lineage movement events among MG, and ES or RJ were observed more frequently than in previous studies, perhaps because this study included more sequences that were sampled close to state borders^6,30^ (**Figure 2B)**.

**Figure 2:**
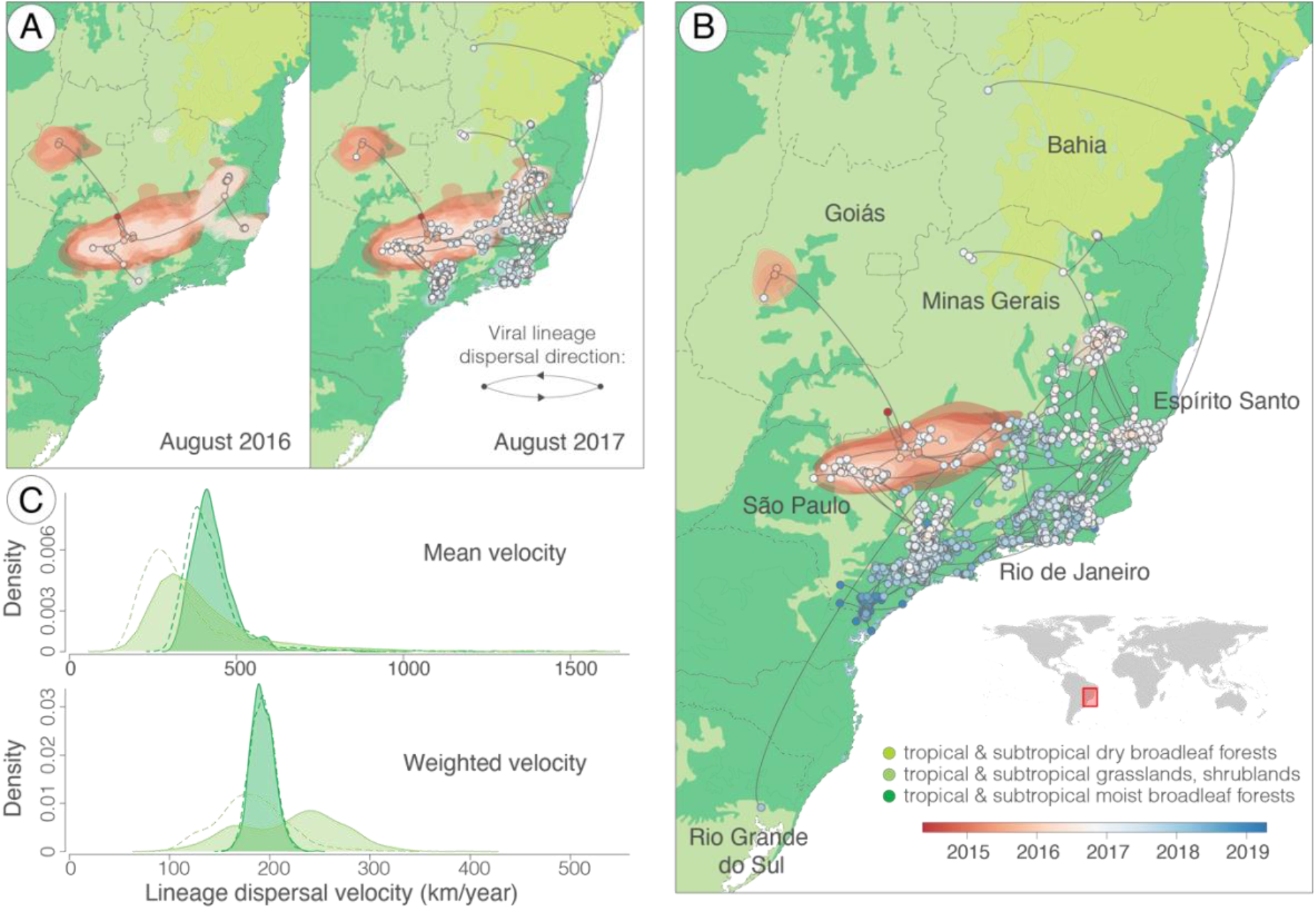
Reconstructed YFV dispersal. **A and B:** maximum clade credibility (MCC) tree branches are mapped according to phylogenetic node and tip locations (circles). Shaded polygons show 80% highest posterior density contours for successive time slices (red to blue scale). State boundaries are shown. Arc curvature direction indicates dispersal direction, as shown in **A. A** shows viral lineage movement until August 2016 and August 2017, and **B** shows all events up until the most recently sequenced genome. Base map colouring shows primary ecoregions. **C:** Density of mean lineage dispersal velocity (i.e., mean of individual branch velocities) (upper) and weighted lineage dispersal velocity (i.e., total estimated distance travelled divided by total time, as represented by the whole tree^10^) (lower), coloured according to ecoregion following **B**. Filled areas show results obtained using genome sequences from all host species, and dashed lines show results obtained from analysis of viruses from non-human species only.

YFV case data shows that patients sometimes acquire infections in a location far from the location of disease notification (**Extended Data Figure 2**), but travel histories were not available for all patients. We therefore estimated the mean lineage dispersal velocity using two different datasets; a dataset of 705 human, NP and mosquito-derived YFV genomes (“full dataset”), and a smaller dataset of 466 sequences from which human-sourced genomes were removed (“non-human dataset”). Analyses of both datasets concluded that virus lineages tended to disperse at 0.6 km/day (95% CI 0.5 – 0.7 km/day), indicating that unrecorded human travel away from infection location did not strongly bias estimates of virus lineage movement velocity in our dataset (**Extended Data Table 1, Extended Data Figure 3**). The mean dispersal value is comparable though slightly slower than that estimated previously from precisely geolocated YFV infections in *Alouatta spp*. in São Paulo state (∼1.4 km/day, with 71% of estimated movements being <1km/day)^38^. Further research would be useful to determine whether this discrepancy arises from inherent methodological differences in estimating dispersal routes, or from faster viral movement in *Alouatta spp*. or São Paulo state compared to across the whole outbreak involving additional NP species.

### Factors associated with YFV dispersal routes

Several factors have been hypothesised to explain variation in the route and rate of YFV dispersal^39^, including the impact of long-distance wind-dispersal of infected mosquitos^40^, altered blood-meal choice of mosquitos in degraded habitats^41^, movement of infected humans, or human-mediated transport of infected animals and vectors^6,40,42^. We used four phylogeographic approaches to test the association of the route and speed of lineage movement with environmental and demographic factors.

We first used a generalised linear model (GLM)^43^ to parameterize transition rates between locations and explicitly test which of 23 different covariates (listed in **Data S1**) were most associated with virus movement rates between those locations. Covariates were identified via a literature review (**Supplementary Note 1**), followed by subsequent removal of highly co-correlated variables. The discrete phylogeographic GLM approach requires that sequences are pre-allocated to groups based on geographic clustering. To check robustness to different clustering, we performed the same analysis for three sets of differently defined discrete locations based on (i) mesoregion boundaries, and on sequence sampling municipalities grouped using (ii) k-means clustering and (iii) hierarchical clustering algorithms (**Figure 3**). Given that the outbreak in Brazil is caused by the sylvatic cycle of YFV^6^, internal phylogeny branches typically represent transmission between NPs, whereas external branches linked directly to tips represent both transmission among NPs, and from NPs to humans. To begin to separate out factors that might differently influence transmission occurring within different hosts, we separately considered the impact of factors on transmission events determined using internal and external branches.

**Figure 3:**
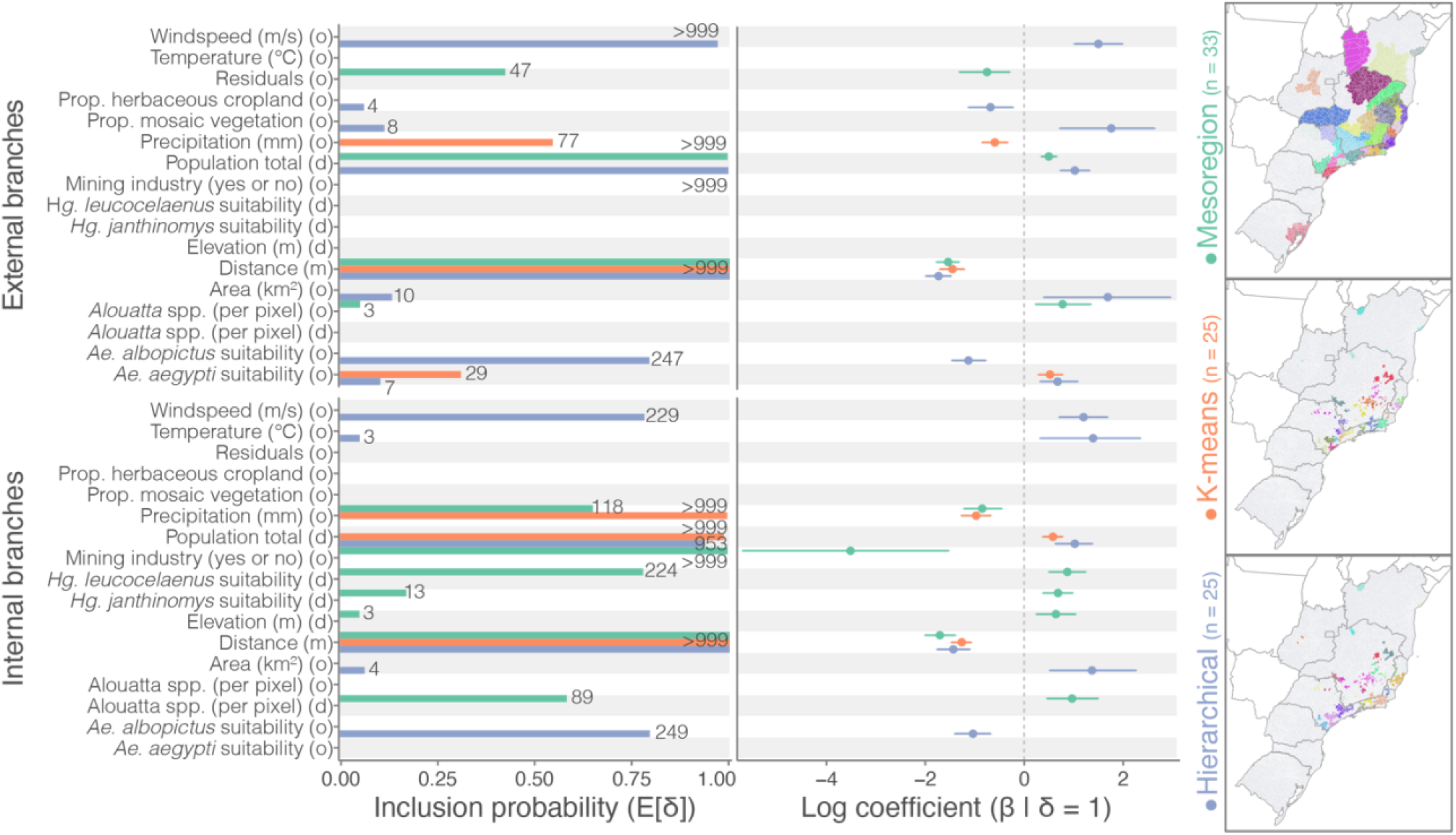
YFV lineage movement drivers. **Left column** shows model inclusion probability for different covariates. Only covariates with appreciable Bayes factor (BF) of >=3 are shown, following^44^. Labels show BF. Labels marked “(o)” and “(d)” indicate separate origin and destination effects for each covariate, respectively. “Prop.” = proportion of land covered by either herbaceous, rainfed crops or >50% mosaic vegetation (ESA CCI class 11 and 40, respectively). Climatic covariates are annual means. **Middle column** shows log coefficient estimates for the covariate conditional on the covariate being included in the GLM. Bars show 95% HPD intervals. Estimates in the upper and lower rows of the left and middle columns are derived from external and internal phylogenetic branches, respectively. Colours in these columns correspond to the clustering algorithm used to group sequences into discrete geographic locations, as shown in the label colours provided in the **right column**. All covariate estimates are provided in **Data S2** and details of sequence geographic grouping in **Data S3**.

In all three spatial groupings, the distance between region centroids was consistently included as a strong predictor of lineage movement for both internal and external branches (inclusion probability of 1, Bayes factor [BF] > 999), indicating that movement events occurred more frequently between nearby locations (**Figure 3**). In contrast, no environmental or demographic variables were identified in all three analyses. Factors identified in only one or two analyses, such as lower suitability for *Aedes aegypti* in the location of origin, may not be robust, or may be differently important when considered at different spatial scales.

To test the impact of environmental covariates on YFV dispersal at a finer spatial resolution, we subsequently used several approaches based on continuous phylogeography. We repeated each analysis using both the full dataset (representing viruses from all hosts) and the non-human dataset (representing viruses from NPs and mosquitos only) to check whether human movement away from the location of infection affected results.

We first conducted an exploratory univariate analysis into whether the locations through which YFV lineages dispersed tended to be characterised by particular environmental conditions. These analyses can be more influenced by sampling effort and pattern than several of the other analyses described here, so represent a description of the environmental context of inferred virus lineage dispersal rather than a robust test of the impact of those conditions on dispersal^45^. Environmental variables that were strongly supported in analysis of the full dataset were always strongly or moderately supported by analysis of the non-human dataset, and vice versa (**Data S4** and **Data S5**). We found that YFV lineages tended to avoid dissemination through (i) areas with high suitability for *Aedes aegypti* mosquitos (BFs = 37.5 and 46.6, for non-human and full datasets respectively); (ii) areas covered exclusively by herbaceous crops (BFs = 18.6 and 89.9); and (iii) areas with higher annual mean temperatures (BFs = 26.8 and 19.0). Instead, YFV lineages circulated preferentially in (i) regions with mosaic vegetation (>50% mosaic tree, shrub and herbaceous cover, and <50% cropland) (BFs = 14.2 and 21.7); (ii) in wetter areas (based on two correlated metrics of high aridity indices, BF = 34.7 and 33.5, and mean annual precipitation, BF = 4 and 3 (correlations provided in **Data S6**)); and (iii) in areas with high density of one or more of six NP genera (*Alouatta, Brachyteles, Callicebus, Callithrix, Leontopithecus, Sapajus*) (BF > 99 for both datasets). Determining whether specific genera are particularly important in supporting YFV spread was not feasible, because host genus range estimates frequently overlap (e.g., **Data S7, Data S8, Data S9**) and because phylogenetic approaches to reconstruct genus-specific transmission yielded conflicting results (**Supplementary Note 2**). We also found that proximity to main roads, intensity of nightlights, urban areas, and human population density were all each independently associated with YFV dispersal (BF > 99 in both datasets). The latter two variables are highly co-correlated (Pearson’s correlation coefficient 0.88, **Data S6**). We consider these findings likely to reflect bias towards NP carcass reporting in more densely human-populated areas, because YFV infections in NPs are detected more frequently in regions with high human population density (**Extended Data Figure 4)** because NP carcasses in these areas are more frequently notified to public health authorities for sampling.

Second, we investigated whether YFV lineages tended to remain within the same ecoregion. Ecoregions are geographically distinct assemblages of species and environmental conditions. The Brazilian study region primarily includes “tropical and subtropical grasslands, shrublands” and “tropical and subtropical moist broadleaf forests” (**Figure 2**)^46^. We found positive support (BF = 8.4 in the full dataset, BF = 6.69 in the non-human dataset) for the hypothesis that YFV lineages tend to avoid migration events between different ecoregions.We also compared the dispersal velocity of lineages circulating in the two primary ecoregions in the study area (**Figure 2C**). Whilst there is a tendency for mean dispersal velocities to be lower in broadleaf forests compared to grasslands and shrublands, this difference is not supported for the full dataset (BF = 2.14), is only substantially supported by the NP dataset (BF = 4.32) and is absent when considering weighted branch dispersal velocity (**Figure 2C**).

Finally, we investigated whether specific environmental factors were associated with differences in the velocity at which YFV lineages spread. Our analyses use a Bayesian approach that assesses the correlation between environmentally scaled distances (*i.e*., geographic distances that are weighted according to the environmental conditions through which lineages pass) and lineage dispersal duration^47,48^. We found strong support (BF = 49 using the non-human dataset, and BF > 99 using the full dataset) that YFV lineages tended to disperse slower in higher elevation areas and faster in higher temperature areas (BF = 15.7 in the non-human dataset, and 8.1 in the full dataset) (**Data S10** and **Data S11)**. The association of elevation and virus dispersal velocity may be being driven in part by lower temperatures at higher elevations (Pearson’s correlation coefficient 0.69, **Data S6**). Overall, only a small proportion of the variability in lineage dispersal velocity could be explained by the environmental heterogeneity in any supported factor within the study area (**Data S10 and Data S11**). Analyses using the full dataset showed a tendency for lineages to disperse faster in areas with high human population density or urban areas (BF = 24 and BF = 15.7, respectively), but slower in areas that were further from main roads (BF >99). These latter results were not replicated however using data from mosquitos and NPs only, and may therefore represent a bias caused by unreported movement of infected humans away from the site of infection prior to notification.

### Environmental factors associated with YFV seasonal dynamics

Identifying factors that explain changes in viral prevalence in the zoonotic reservoir over time is important to predict when YFV spillover risk to humans may be highest. We used a GLM extension of a skygrid coalescent approach^49,50^ to quantify the association between virus effective population size and environmental covariates. Effective population size is a population genetic parameter related to number of infected individuals, though often non-linearly^51^. We first used environmental covariates for the region most affected by YFV, covering the states of MG, SP, RJ and ES (**Figure 2**, “state-based analyses”). We also included two metrics of agricultural seasonality that had been identified through a previous modelling study as being associated with the number of reported YFV cases^52^.

The number of confirmed YFV-infected NPs was the best predictor of virus effective population size; this finding is expected because YFV infection counts are an outcome of the virus infection process (inclusion probability of 0.52, regression coefficient of 0.71) (**Figure 4**). Sea surface temperature (SST) anomalies in the Niño 3.4 Region (5N–5S, 120W–170W) are commonly used indicators for characterising the El Niño Southern Oscillation^53^. SST was weakly associated with virus population size (**Figure 4**) but could be considered a driver of other included climatic variables and was therefore removed, along with YFV infection counts, in a second analysis. Higher temperature was consistently identified as a predictor of greater virus effective population size and was the best predictor after number of YFV infected NPs and SST anomalies were removed from the analyses (**Figure 4A and Figure 4B**). To assess the robustness of our findings, we performed an additional skygrid-GLM analysis that was parameterised using environmental data from only those areas estimated to be experiencing YFV circulation in each time window. Whilst the strength of association between temperature and virus effective population size was not meaningfully affected (inclusion probabilities 0.51 and 0.49), confirmed YFV infections from regions with ongoing circulation were more closely associated with virus effective population size dynamics than YFV infections from the four most affected states (inclusion probabilities 0.945 and 0.055, respectively). These analyses confirmed that infection counts and mean temperature were the best predictors of virus effective population size, indicating that our results are likely robust to the spatial area for which environmental data was extracted (**Figure 4)**.

**Figure 4:**
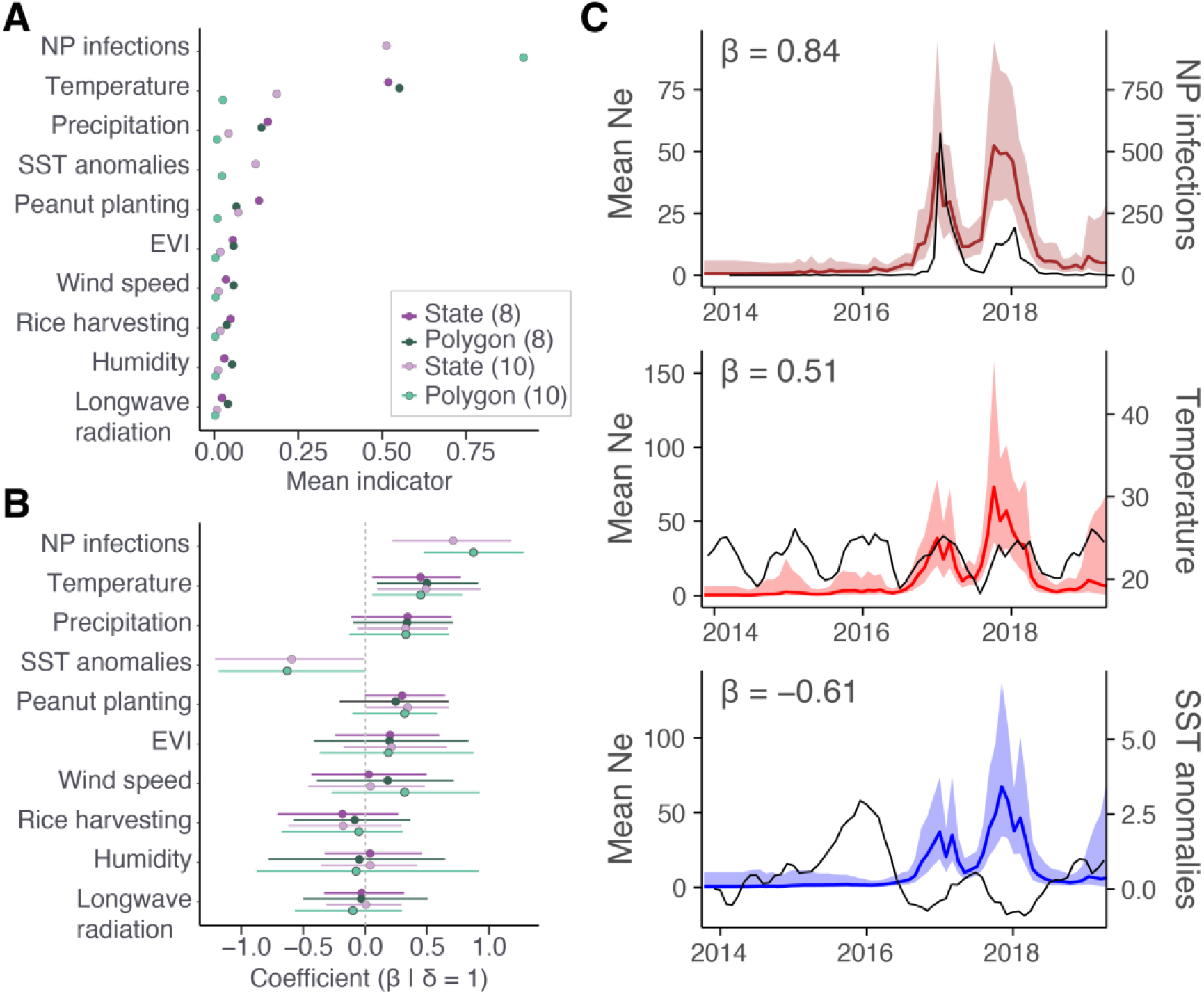
Factors associated with YFV effective population size. **A:** mean of indicators for each variable. Legend shows four different analyses (“state-based” with 8 or 10 covariates, and “polygon-based” with 8 or 10 covariates). **B**: Regression coefficients (β) when the corresponding bimodal indicator is 1, legend as in **A. C**: associations between mean viral effective population size (Ne) and time-dependent covariates (state-based analyses, see **Extended Data Figure 5A and B** for polygon-based analyses); number of confirmed YFV infections in NPs, mean annual temperature (°C) and sea-surface temperature anomalies. Coloured lines show mean Ne estimated from viral sequence and covariate data and ribbons show the 95% HPD. Black lines show the mean of covariates. Covariate values were extracted per month for the region containing the federal states of SP, ES, RJ and MG.

Variables that best predict effective population size in the following month may be most useful to help nowcast outbreak dynamics in the following weeks, and thereby enable public health preparedness or escalation of preventative vaccination campaigns. We therefore conducted a skygrid-GLM analysis to estimate which variable best predicts effective population size estimated in the following month. The proportion of rice farms conducting harvesting was inversely associated with virus effective population size one month after measurement (**Extended Data Figure 5C–E**). We consider it very unlikely that this relationship is directly causal, but it is possible that increases in harvesting may capture a joint effect of reducing temperature and rainfall, and therefore track reduced suitability for arbovirus transmission better than either climatic variable alone.

### Mechanism of persistence between inter-epidemic periods

Remarkably little is known about how YFV is maintained during cool, dry seasons with poor suitability for mosquito-borne virus transmission, yet this is crucial to understand whether transmission can be interrupted before large epidemics develop during warmer and wetter seasons. Several hypotheses have been put forward, including virus re-introduction from locations with higher suitability, and persistence of vertically transmitted (transovarial) virus within unhatched mosquito eggs^54^. Vertical transmission of YFV has been demonstrated in laboratory settings for the urban mosquito *Aedes aegpyti*^34 55,56^, and is plausible for the primary sylvatic YFV mosquito vector, *Haemagogus spp*., because eggs can remain viable for several months during dry periods^57^. However, it is unknown whether vertical transmission contributes substantially to epidemiological dynamics in natural sylvatic outbreaks. We hypothesized that widespread and durable persistence of YFV in mosquito eggs during seasons with low suitability for YFV transmission would be associated with a lower evolutionary rate during those seasons, because the presumably non-replicating (or minimally replicating) virus would be held in evolutionary stasis. To investigate how YFV persists between epidemic waves, we performed a phylogenetic analysis with a time-heterogeneous epoch substitution model^58^.

We estimate that YFV evolves 1.7 times faster during the six-month annual period in which YFV transmission is highest, compared to the six-month period when transmission is lowest (**Figure 5A, Data S12**). More sequences were available from high transmission periods, and therefore the relative number of tree branches that are external branches is likely higher during these periods. To ensure that the higher evolutionary rate was not driven by the more frequent accumulation of deleterious mutations in external branches^59^, we conducted further epoch analyses considering rates calculated using only internal branches. We also ran separate analyses focused on internal branches within the two major clades that primarily circulated in São Paulo (YFVSP), and that primarily circulated in Minas Gerais, Espírito Santo and Rio de Janeiro (YFVMG/ES/RJ) (marked on **Figure 1**). Our results remain robust (**Figure 5B**). We subsequently introduced a lag in our epoch definitions, such that epoch transitions occurred in the peak and troughs of confirmed YFV infections. As expected, there was no difference between virus evolutionary rates in these time-shifted epochs (**Figure 5**).

**Figure 5:**
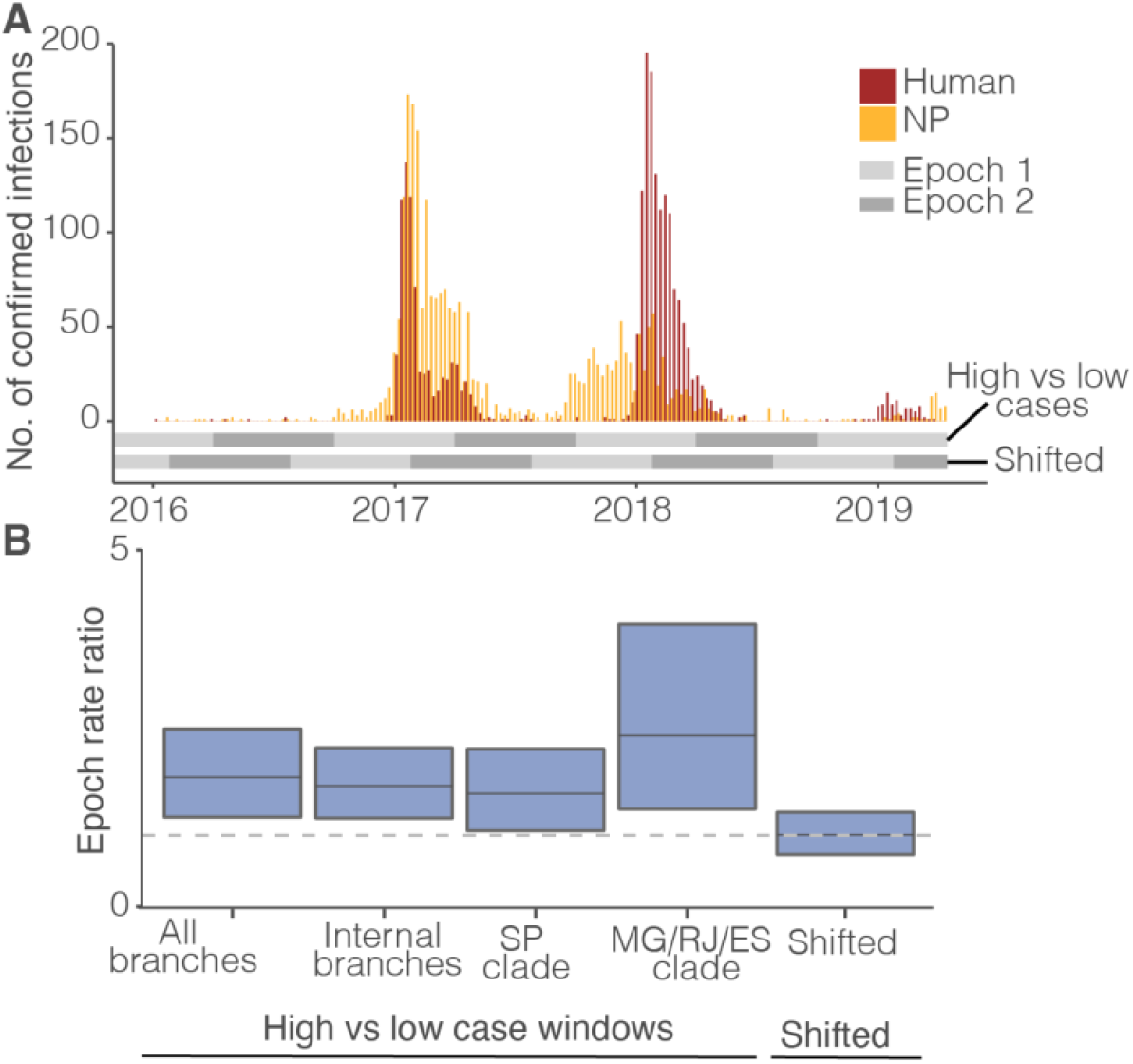
Seasonally varying evolutionary rate. **A**: Vertical bars represent the number of confirmed infected humans (brown) and NPs (yellow), and horizontal lines represent epochs used in different analyses. **B:** Ratio of the evolutionary rate in epoch 1 to the rate in epoch 2 at each sampled MCMC step for different analyses. Boxplots define 95% HPD intervals of the ratio, with the median marked horizontally. The horizontal dotted line indicates the ratio of 1. “All branches” shows results when internal and external branches in each epoch are specified by the same rate, “internal branches” shows results from internal branches only when internal and external branches are specified using different rates, “SP clade” and “MG/RJ/ES clade” show results focusing on each major phylogenetic clade, and “shifted” shows results when internal and external branches are specified separately and the epoch windows are lagged by 3-months.

Together, our results indicate that the evolutionary rate of YFV is higher in periods of high transmission than in periods of low transmission. Our findings could be explained by at least two processes. Firstly, hatching rates of sylvatic *Haemagogus* mosquitos are higher following frequent rainfall^57^. If YFV is able to persist in evolutionary stasis in sylvatic mosquito eggs without undergoing replication^55,56^, then a shorter duration prior to hatching may result in a faster evolutionary rate in warm, wet seasons. Slower rates of virus dispersal in winter have been previously reported based on case data^38^, which would be consistent with YFV persistence in (stationary) mosquito eggs. We did not detect a strong seasonal difference in dispersal velocity during continuous phylogeographic analysis described above (**Extended Data Figure 4**), although explicitly testing this was not our intention and therefore the chosen method would likely have been underpowered relative to possible alternative epoch-based model specifications. Further extending this phylogeographic epoch model to formally test whether branch velocities are lower in winter would be valuable. Secondly, the extrinsic incubation time for YFV in mosquitos is believed to be temperature-dependent^60^. Whilst analyses on sylvatic mosquito vectors are lacking, the 5°C oscillation in temperature observed in the Brazilian states considered here (shown in middle panel, **Figure 4C)** is predicted to increase the incubation time in *Aedes aegypti* by approximately 30%^60^. Assuming that incubation time is negatively correlated with the replicative rate of the virus, a decrease in incubation time could partly explain the higher evolutionary rate observed in warmer, high transmission seasons (**Figure 5**).

Southeast Brazil is experiencing a trend towards higher temperatures^61^ and higher frequency, heavier rainfall^62^. It is important to experimentally validate whether these climatic changes may indeed cause higher virus evolutionary rates, and study whether this could contribute to increased risk of YFV (or other arboviruses) switching from a sylvatic to an urban cycle.

## Conclusion

We report a comprehensive genomic analysis of YFV spread in Brazil from 2016 to 2019. We show that greater YFV spread is strongly associated with decreased geographic distance between locations. Environmental factors, including high density of NPs, wetter areas, and higher temperature, were associated with increased speed and/or differences in the route of geographic spread. However, perhaps because the study region is comparatively environmentally homogeneous relative to variation at a continental scale, detected effects were relatively small. In contrast, our findings show that higher temperatures are very strongly associated with increased virus effective population size. Seasons with higher virus population sizes are also associated with more rapid virus evolution. The current rate of global warming is unprecedented in recent human history^63^ and may lead to increased YFV transmission in some locations^64^. International strategies to vaccinate human populations must therefore be maintained and enhanced to anticipate a scenario in which yellow fever outbreaks could become larger, faster spreading, and with greater potential for acquiring virus genomic changes. Multidisciplinary approaches to understand how YFV epidemiology may change with increased urbanisation, environmental and climate change are important to complement human immunisation campaigns, as outlined in WHO’s Eliminating Yellow Fever Epidemics (EYE) Strategy^65^. Our study underscores the urgent need for focusing on zoonotic virus transmission at the human-animal-environment interface and the high potential of phylodynamic approaches in explaining the origins and dynamics of zoonotic disease.

## Supporting information

Supplementary Information (contains Supplementary Notes 1 - 9, Extended Data Figures 1 - 10, Extended Data Table 1)

## Data Availability

All newly generated sequences are deposited on GenBank, with accession numbers ON022238 - ON022746. XML code for phylodynamic analyses is available in Data S19.

## Materials and Methods

### Epidemiological data

We obtained anonymised data on human and neotropical primate (NP) confirmed yellow fever virus infections (YFV) via the Brazilian Ministry of Health from the National Notifiable Disease System (SINAN; Sistema de Informação de Agravos de Notificação). The Ministry of Health considered confirmed infections to be those positive via RT-qPCR, immunohistochemistry and/or IgM ELISAs.

### Sample collection

Our research represents a co-ordinated effort by four institutes; Fundação Oswaldo Cruz in Rio de Janeiro (Fiocruz), Fundação Ezequiel Dias in Minas Gerais (Funed), Instituto Adolf Lutz in São Paulo (IAL), and Instituto de Medicina Tropical (IMT), Universidade de São Paulo, São Paulo (IMT-USP). Samples were collected by these institutes during routine YFV surveillance from patients attending public health services, or from NP carcasses collected following public reporting. Mosquito samples were collected by IAL through active surveillance in risk areas.

### Ethical statements

Research at Fiocruz and Funed was supported by the Pan American Health Organization (PAHO) and the Brazilian Ministry of Health as part of arboviral surveillance efforts according to Resolution 510/2016 of the National Ethical Committee for Research (Comissão Nacional de Ética em Pesquisa; CONEP). YFV genomic diagnostics on residual human diagnostic samples was approved by the Ethics Committee of the Oswaldo Cruz Institute (CAAE90249218.6.1001.54248).

Genomic surveillance of NP carcasses by IAL was approved by the IAL Ethics Committee for Animal Use in Research (0135D/2012 and 020G/2014). Research on residual, anonymised human samples obtained by IAL was supported by the Brazilian Ministry of Health as part of arboviral genomic surveillance efforts according to Resolution 510/2016 of CONEP (National Ethical Committee for Research, Ministry of Health).

Research on residual, anonymised human samples collected at Hospital das Clínicas and sequenced at IMT-USP was approved by the HC-FMUSP Ethics in Research Committee (#2.669.963, #3.258.615, and #3.371.745).

### Virus genomic sequencing

We extracted total RNA using commercially available kits, and tested samples for the presence of YFV RNA using previously published RT-qPCR methods^66–70^. The choice of kits and protocols varied according to routine procedures for that sample type and institution (detailed in **Supplementary Note 3**).

We generated most YFV genome sequences (n = 491) using the Oxford Nanopore Technologies (ONT) MinION, following previously validated protocols^6,71^. In brief, we generated cDNA using random hexamers before amplifying YFV cDNA using the multiplex PCR described in^6^. We sequenced purified PCR products only where the dsDNA concentration measured using a Qubit 3.0 or 4.0 fluorometer (Thermofisher) was greater than 4 ng/uL, indicative of possible successful amplification. We carried 10 ng of each sample forward into sequencing libraries that each contained 23 samples and 1 negative control. We prepared libraries according to the ‘Baseline’ protocols Steps 16–22 in dx.doi.org/10.17504/protocols.io.bdp7i5rn^72^, but with minor differences described in **Supplementary Note 4**. We loaded 30 ng of library onto ONT FLO-MIN106 flow cells, and visually monitored the output of sequencing using Rampart (https://github.com/artic-network/rampart).

We generated consensus sequences using protocols published by the ARTIC network (https://github.com/artic-network) ^6,28,71,72^. In brief, we basecalled raw fast5 data using Guppy version 3.0.3 (ONT). We demultiplexed reads and trimmed sequences using Porechop version 0.2.3 (https://github.com.rrwick/Porechop). We used bwa^73^ to map reads to a reference genome (GenBank accession number JF912190). We identified variants from the reference genome using nanopolish version 0.11.1 variant caller^74^, replacing sites with <20x coverage with an ambiguous base (N).

We sequenced 7 samples using an Illumina MiSeq according to the protocol detailed in ^6^. The method used to generate each genome is shown in **Data S13**. We also report accession numbers for those sequences previously generated as part of Kallas et al^69^.

### Compilation of genetic datasets

We downloaded YFV genome sequences from GenBank (19^th^ August 2020). We excluded sequences between 2017–2020 that had not yet been included in a publication by the submitting authors (n = 30). We genotyped sequences using a YFV genotyping tool ^6^ and retained YFV SA1 genotype genomes. We randomly removed duplicate entries from the same patient where identified.

We aligned YFV genomes obtained from GenBank with genomes generated in this study using MAFFT v.7 ^75^, and checked the alignment manually to remove any sequences suspected of containing sequencing errors. We trimmed to YFV coding regions and removed sequences shorter than 70% of the coding sequence length (<7166bp).

We confirmed the monophyly of all sequences sampled during the recent Brazilian outbreak using a maximum likelihood tree estimated in RAXML-NG^76^. We used a general time-reversible model (selected using ModelTest-NG^77^) and a discretised gamma distribution to model among-site rate variation along with a proportion of invariant sites (GTR + Γ4 +I). We retained sequences within the monophyletic outbreak clade that had minimum necessary metadata (year and municipality of sampling) (n = 705, of which 485 were generated here) (**Data S13**).

To confirm the adequacy of our dataset for molecular clock phylogeographic analyses, we checked that our data had appropriate temporal signal using TempEst v1.5.3^78^ and confirmed that our sequence sampling was not highly biased relative to counts of confirmed YFV infections (**Supplementary Note 5**).

### Clustering of sequences into discrete groups for generalised linear model (GLM) phylogeographic analyses

We assigned genetic sequences to discrete groups based on their geographic location of sampling to enable analysis within a discrete phylogeographic framework. We explored three different groupings to determine whether chosen groupings impacted the significance of association of covariates with virus dispersal (grouping maps shown on **Figure 3**). The chosen groupings were based on (i) mesoregion (as defined by the Brazilian Institute of Geography and Statistics (IBGE) ^79^) (n = 33), (ii) k-means clustering (n = 25), and (iii) hierarchical clustering algorithms (n = 25). Additional methodology is provided in **Supplementary Note 6**.

### GLM covariate selection and transformation

We used a phylogeographic GLM^43^ to determine factors associated with virus movement between different locations. We considered explanatory covariates that were identified through a literature review (PubMed search on 29^th^ July 2020), as well as several variables that were not previously considered. Identified covariates include climatological (e.g., rainfall, temperature), ecological (e.g., mosquito and host distribution), physical (e.g., altitude) and anthropogenic (e.g., vaccination, deforestation) factors, and are presented in **Supplementary Note 1**.

We processed variables from geospatial datasets from both vector and raster data sources (as presented in **Data S14**) to calculate summary statistics for spatial units (raw summary statistics are given in **Data S15**). We chose data sources with temporal coverage of 2015–2019 inclusive as far as possible, corresponding to the timing of the outbreak studied here. We typically considered long-term mean, minimum and maximum values for the discrete geographic region (**Data S14**). We clipped all raster datasets to the spatial extent of the study area, whilst maintaining the native spatial resolution of each dataset. We calculated summary statistics for (i) mesoregions, and groups of municipalities, clustered using (ii) hierarchical clustering and (iii) a k-means approach, as defined above. For each unit we also calculated the geographic area, the distance between pairs of centroids, whether units shared a boundary, and the length of the shared boundary. Pre-processing of the input datasets was necessary prior to calculating summary statistics of the spatial units for each variable, as described in **Supplementary Note 7**. Data describing two further variables were only available as summaries at the municipality level (presence of substantial mining industry and vaccination coverage; data sources in **Data S14**). Details of data processing are described in **Supplementary Note 7**.

The phylogeographic GLM model parameterises the log of the between-location transition rates as a log linear function of the predictor ^43^. We therefore log-transformed continuous predictors, and standardised values for each covariate to grant predictors equal variance. Values representing presence or absence were encoded as 1 and 0 without further transformation. We checked for collinearity between variables using the *vifcor* function from the R package *usdm*^80^ (values presented in **Data S7, Data S8** and **Data S9**). We typically removed all but one of each group of variables that were collinear with a correlation coefficient of over 0.75. Variables selected for use in phylogeographic GLM are shown in **Data S1**.

### Discrete trait phylogeographic GLM

We used a phylogeographic GLM^43^ implemented in BEAST 1.10.5^81^ to determine factors that influenced the dispersal of YFV between locations. Each sequence was first tagged with a location of sampling for each of the three clustering algorithms described above. The GLM parameterises the movement of virus lineages between pairs of these locations as a log-linear function of potential predictors using a coefficient that quantifies the effect size of that predictor, and a binary indicator variable that allows the predictor to be included or excluded from the model. We used a prior distribution on these indicators such that there is a 50% prior probability that no predictors are included in the model. We specified separate predictors based on both the conditions at the origin and the destination in order to capture both ‘push’ and ‘pull’ influences. We tested for the impact of sampling bias by also including a predictor based on the residuals of the regression of genetic sequence sample size against all confirmed YFV infections in humans and NPs for both the origin and destination.

All molecular clock phylogenetic analyses were completed in BEAST 1.10.5^81^ using the BEAGLE library v3.2.0 to improve computational speed^82,83^. We always combined at least two independent chains to confirm convergence at the same point and removed at least the first 10 % of each chain as burn-in in each BEAST run (and sometimes longer where convergence was not reached by 10 % as identified by visual inspection using Tracer v1.7.1.). Markov chain Monte Carlo (MCMC) algorithms were run for up to 1,000,000,000 steps but typically fewer, depending on the complexity of the analysis.

We used a subsampled set of 1000 empirical posterior trees generated in BEAST^81^ as input to the phylogeographic GLM. We generated this tree set using the HKY substitution model with gamma-distributed rate heterogeneity, an uncorrelated lognormally distributed relaxed clock model^84^, and a skygrid coalescent model with 60 population sizes. The number and spacing of population size parameters was chosen to allow for one grid point per calendar month between the youngest tip in 2019, and the estimated most recent common ancestor in 2014)^50^. Due to excessively long chain completion times, the HKY model was chosen to improve computational speed and convergence relative to the GTR model identified through ModelTest-NG.

We used the phylogeographic GLM to determine whether different factors were important for lineage dispersal events represented by internal branches (primarily representing transmission in the sylvatic reservoir) and those represented by external branches (representing both transmission within the sylvatic reservoir, and probable dead-end spillovers to humans). We achieved this using a branch partitioning model in which the association of different GLM covariates with lineage dispersals is predicted independently for internal and external branches^85^. Highest posterior density intervals were calculated using the package *coda* version 0.19.4^86^ in R version 4.0.5^87^ within RStudio version 1.4.1106^88^, and data plotted using *ggplot2* version 3.3.2^89^.

### Covariates for skygrid analyses

To determine how seasonally-varying factors are associated with virus effective population size over time, we used a GLM extension of the skygrid coalescent model (henceforth “skygrid-GLM”) to test for associations between different covariates and the estimated virus effective population size^49,50^. Chosen covariates included confirmed infections in NPs reported to SINAN, and climatic and agricultural data (**Data S16)**. Details of data extraction are described in **Supplementary Note 8**.

We extracted data for the region that represented the focus of the epizootic in Brazil in two different ways. We first aggregated climatic and infection data across the four states with the highest reported infection burden: São Paulo (SP), Rio de Janeiro (RJ), Minas Gerais (MG) and Espírito Santo (ES). These states reported 88% of all confirmed NP infections and 98% of all confirmed human YF cases in Brazil in the time frame shown on **Extended Data Figure 6**.

We extracted values for the selected region using a shape file of Brazilian states obtained from the Brazilian Institute of Geography and Statistics (IBGE)^79^ and trimmed to Brazilian states of interest. We processed the data using the *raster* package, *extract* function in R^90^. In addition to climatic data, we also used two covariates representing seasonal variation in agriculture, choosing the two predictors that have been previously shown to be most explanatory of YF case counts in Brazil (rice harvesting and peanut planting). We obtained data from a previous study^91^, and transformed them to represent the proportion of all peanut/rice farms in an area that were conducting each activity per month. Unlike the climatic data we use, these data were only available for one representative year, and therefore data from this year were repeated annually to represent multiple years. We log-transformed and standardised all covariate values.

Secondly, we used an approach informed by the results of continuous phylogeography to extract a second set of covariate values that could account for the shifting epicentre(s) of the YFV outbreak over time. Specifically, we used the R package “seraphim”^92^ to extract the minimum convex polygons surrounding all inferred phylogenetic node locations for 5 month sliding windows. 95% highest posterior density polygons were defined using 100 posterior trees. This polygon was used to crop rasters from which climatic data were extracted as described above, or to select municipalities from which confirmed YFV infection counts were aggregated. All confirmed infections in municipalities crossed by polygon edges were included, as more precise locations below the municipality level were unavailable.

We used the *vifcor* function from the R package *usdm*^80^ to identify climatic variables that were highly collinear, and remove these before analysis. Correlations between variables for both analyses are presented in **Data S17** and **Data S18**. Covariates that we chose for inclusion are listed in **Data S16**.

### Skygrid-GLM

We used the skygrid-GLM approach implemented in BEAST 1.10.4 ^81^ to investigate the impact of time-varying environmental covariates on virus effective population size. We used monthly skygrid intervals. We completed runs using different covariate sets listed in **Data S16**, based on variables selected before the analysis to avoid multicollinearity. When predictors were robustly associated with changes in effective population size, we used a univariate skygrid-GLM to more accurately estimate the correlation coefficient (**Figure 4C** and **Extended Data Figure 5A and Extended Data Figure 5B**).

For the two best associated covariates (number of confirmed YFV infections in NPs, and annual mean temperature), we used skygrid-GLMs with two indicators to directly compare whether data obtained from state boundaries, or data obtained from within polygons identified during continuous phylogeography, better explained the dynamics of virus population size.

We additionally considered whether inclusion of a one-month lag period between the covariate and the virus effective population size was still able to identify covariates with significant association. Specifically, we considered whether any of 9 climatic and environmental covariates were predictive of effective population size one month later (**Data S16)**. As our previous results were highly consistent between state-boundary-based and polygon-based analyses, we only conducted these analyses on the covariate dataset extracted using state boundaries.

### Continuous phylogeography

We used a continuous trait phylogeographic approach to reconstruct the dispersal history of YFV lineages in continuous space. Specifically, we used a relaxed random walk model implemented in BEAST 1.10.5^81^, with a gamma distribution to model the among-branch heterogeneity in dispersal velocity^93,94^. For this analysis, the nucleotide substitution process was modelled according to a GTR+G4 parameterisation^95^, branch-specific evolutionary rates were modelled according to a relaxed molecular clock with an underlying log-normal distribution^84^, and a flexible nonparametric skygrid coalescent model was used as the tree topology prior^50^. The MCMC was ran for >4×10^12^ iterations while sampling every 1×10^6^ iterations and discarding the first 10% of trees sampled from the posterior distribution as burn-in. Convergence and mixing were examined using the program Tracer 1.7^96^, assessing that estimated sampling size (ESS) values associated with estimated parameters were all >200, and TreeAnnotator 1.10.5^81^ was used to identify and annotate the maximum clade credibility (MCC) tree. Spatio-temporal information embedded within trees were extracted using the R package “seraphim”^48,92^. This package was also used to map the resulting continuous phylogeographic reconstruction and to estimate two dispersal statistics: the mean lineage dispersal velocity and weighted lineage dispersal velocity estimates. The former is calculated as the mean velocity of all individual branches, whereas the latter is calculated as the sum of the total great circle distance moved during every branch, divided by the sum of time represented by every branch ^10^.

YFV samples were all associated only with a municipality of origin. Because the RRW model does not allow different sampled sequences to be associated with identical geographic coordinates, we randomly drew, for each sequence, a random sampling point within the administrative polygon of its municipality of origin. We use this approach as an alternative to the more standard procedure consisting in considering the centroid point of the administrative polygon of origin and adding a restricted amount of noise to duplicated identical sampling coordinates. The latter procedure could lead to a situation in which the uniform random noise assigns a sampling location that falls outside the administrative area of origin of the considered sampled sequence^97^.

We used three different, previously described^10,45,47,97^ approaches implemented in the R package “seraphim” ^3,79^ to test the impact of different environmental covariates on the dispersal of YFV lineages. For each approach, we performed analyses based on the full dataset of 705 SA1 YFV genomes from humans, NPs and mosquitos, and also analyses based on an alignment subset containing only the 466 SA1 YFV genomes from NPs and mosquitos. All approaches use an approximation of Bayes factor (BF) support^10,98^ to test whether dispersal frequency, position, or velocity of lineage dispersal is significantly different from that of a null dispersal model, *i.e*. a dispersal model in which the environmental factors did not impact or constraint the dispersal dynamic of YFV lineages. Note that possible maximum value of estimated BFs is artificially limited here by the number of samples in the posterior distribution. We therefore report Bayes factors as >99 where relevant, though the true BF will often be much higher.

We first considered whether YFV lineage circulation tends to avoid switching between different ecoregions, including “tropical and subtropical moist broadleaf forests” and “tropical & subtropical grasslands, savannas and shrublands”^97^. Secondly, we investigated whether YFV lineages tended to preferentially circulate within or avoid areas with particular environmental characteristics^45^. Here, we used univariate analyses to consider the environmental characteristics listed in **Data S14**, some of which which had been previously used for discrete trait GLMs.

Finally, we considered whether YFV lineage dispersal events were made faster (“conducted”) or slower (“resisted”) by each environmental variable in **Data S14**. We used the Circuitscape path model^99^ to calculate environmentally-scaled distances (spatial distances that are weighted according to values of the environment), an algorithm that accommodates uncertainty in the travel route. Further brief description of these methods is given in **Supplementary Note 9**.

### Time-varying evolutionary rate

To investigate whether the evolutionary rate of YFV changed seasonally, we used a phylogenetic epoch model with two repeating seasons^58^. This model allows different strict clock substitution rates to be inferred in different epochs. We initially defined the two epochs as April to September, and October to March, of every year. These seasons correspond to the low and high transmission seasons for YFV as defined by observed infection counts (**Extended Data Figure 7**). Whilst we did not distinguish between internal and external branches in an initial analysis, in all subsequent analyses we specified separate rates on external branches to remove those as a potential confounder. We performed two analyses to confirm that our results were robust, as follows. To establish that epoch rate changes were not driven by different substitution rates in different lineages or locations of transmission, we considered two subsamples of our data, removing first the major clade YFVSP, and second the major clade YFVMG/ES/RJ (**Figure 1**). As a validation for our specific epoch inference approach, we also separately introduced an approximately three-month lag to our previously defined epochs, such that each epoch included parts of both high and low transmission seasons (**Extended Data Figure 7**); specifically, we chose the six-month window that equally partitioned the proportion of infections observed in those windows (**Extended Data Figure 7**). We hypothesised that this would result in equal evolutionary rates in each epoch.

## Data availability statement

All newly generated sequences are deposited on GenBank, with accession numbers ON022238 – ON022746. XML code for phylodynamic analyses is available in **Data S19**.

## Acknowledgments

This work was supported by a Medical Research Council-São Paulo Research Foundation (FAPESP) CADDE partnership award (MR/S0195/1 and FAPESP 18/14389-0) (https://caddecentre.org). SCH was supported by a Sir Henry Wellcome Postdoctoral Fellowship (220414/Z/20/Z). IMC was supported by FAPESP Doctoral Fellowship (2018/17176-8). SD acknowledges support from the Fonds National de la Recherche Scientifique (F.R.S.-FNRS, Belgium; grant n°F.4515.22), from the Research Foundation - Flanders (Fonds voor Wetenschappelijk Onderzoek-Vlaanderen, FWO, Belgium; grant n°G098321N), and from the European Union Horizon 2020 project MOOD (grant agreement n°874850).The Flavivirus Laboratory at Fiocruz is grateful for the support received from Fundação de Amparo à Pesquisa do Estado do Rio de Janeiro /Faperj (grant number E-26/202.930/2016) and by the Coordenação de Vigilância em Saúde e Laboratórios de Referência da Fundação Oswaldo Cruz / Ministério da Saúde do Brasil. GRWW was funded by the MOOD H2020 Project 874850. OJB was supported by a UK Medical Research Council Career Development Award (MR/V031112/1). AEZ and OGP are supported by The Oxford Martin School Programme on Pandemic Genomics (<https://www.oxfordmartin.ox.ac.uk/pandemic-genomics/).> GB acknowledges support from the Internal Funds KU Leuven (Grant No. C14/18/094) and the Research Foundation—Flanders (“Fonds voor Wetenschappelijk Onderzoek—Vlaanderen,” G0E1420N, G098321N). PL acknowledges the support of the Wellcome Trust (Collaborators Award 206298/Z/17/Z – ARTIC network), the European Research Council (grant agreement no. 725422 – ReservoirDOCS) and NIH grant R01AI153044. NRF was supported by a Sir Henry Dale Wellcome Trust Fellowship (204311/Z/16/Z). We also acknowledge support from Oxford Nanopore Technologies for a donation of sequencing reagents. We thank those involved in co-ordinating and conducting yellow fever virus surveillance activities, including Dr. Maria de Fátima Domingos, Sueli Yassumaro Diaz, Simone Lucheta Reginatto, Renata Caporalle Mayo, Sirle Abdo Salloum Scandar, Dr. Lilian Rodas Colebrusco, Ivete da Rocha Anjolete and Raquel Cristina Noronha Silva (SUCEN). For the purpose of open access, the author has applied a CC BY public copyright licence to any Author Accepted Manuscript version arising from this submission.

## Author Contributions

SCH led choice and implementation of analyses and wrote the manuscript. SCH and NRF led conceptualisation of the study. SCH, IMC, PS, TA, MG, JGdJ, FB, AAF, FI, MC, and RSA conducted virus genomic sequencing. IMC, PS, TA, MG, JGdJ, FB, AAF, JX, ACdC, EM, ASSL, EGK, FI, MC, RS, LS, and RSA performed intensive disease surveillance, including diagnostic testing and detailed data curation. SCH, SD, CHW, GB, PL and MAS contributed to phylodynamic analyses. SCH and SD generated all figures. JTdD and LFM performed entomological collections, neotropical primate sample collections and georeferencing. RMT and RMTdM identified mosquito samples. AA and JAMR contributed to coordinating YFV surveillance activities and response. JT, FRRM and SCH adapted virus genomic assembly pipelines and provided bioinformatic support. SL, FC, HRC, OP, NTG, GRWW, PSP, AdSS and AH extracted and processed spatiotemporal covariate data. AEZ helped with statistics. JPM, OJB, AJT, LCJA, RSA, OGP, FI, MSC, AMBdF, ECS and NRF provided supervisory support. FI, MSC, AMBdF, ECS and NRF contributed equally to funding, design and implementation of the study. All authors contributed to review and editing of the manuscript.

## Additional Information

**Supplementary Information** is available for this paper. Correspondence and requests for materials should be addressed to the corresponding authors.

