## Supplementary Information (contains Supplementary Notes 1 - 9, Extended Data Figures 1 - 10, Extended Data Table 1) for "Climate and land-use shape the spread of zoonotic yellow fever virus"

**Supplementary Notes for "Climate and land-use shape the spread of zoonotic yellow fever virus"**

**This PDF file includes:**

Supplementary Notes 1 – 9

Extended Data Figures 1 – 10:

Extended Data Table 1

**Other Supplementary Materials for this manuscript include the following as separate files:**

Data S1 to S19

**Supplementary Note 1**: **Environmental and demographic covariates found in PubMed literature review.** Search terms "(yellow fever virus OR yellow fever) AND (Brazil OR Argentina OR Peru OR Colombia OR Bolivia OR Venezuela OR Chile OR Paraguay OR Ecuador OR Guyana OR Uruguay OR Suriname OR French Guiana OR Panama OR Trinidad and Tobago OR South America)", 29th July 2020. We scanned titles of papers to identify papers focused on statistical modelling or epidemiological identification of factors that affected yellow fever virus presence or transmission. Other variables not considered in other studies, but that were included here, are listed.

| **Type and reason** | **Variables** | **Citation** |
|---|---|---|
| Climate - wind<br><br>*Wind may influence the dispersal of the mosquito vector – areas with higher wind speeds may help disperse vectors.* | Mean / maximum / minimum wind speed / seasonality of wind speed (average variation at each pixel). | [1] |
| Climate - rainfall, humidity, and water vapour pressure<br><br>*Yellow fever mosquito vectors occur more frequently during rainy season – higher rainfall areas may be more likely to be sites of persistence or origin* | Annual mean rainfall | [1–5] |
| | Aridity index | [1] |
| | Average hourly rainfall | [6] |
| | Maximum / average rainfall in wettest month | [1,2] |
| | Minimum / average rainfall in driest month | [1,2] |
| | Mean rainfall in the wettest/ warmest/ coldest/ driest/ most humid quarter | [1] |
| | Mean tasselled cap wetness (surface moisture) | [7] |
| | Potential evapotranspiration (PET) | [1] |
| | Seasonality in rainfall | [1] |
| | Minimum / maximum / annual mean / seasonality in water vapour pressure | [1] |
| | Topographic humidity index | [1] |

| | | |
|---|---|---|
| Climate - solar radiation<br><br>*May drive humidity and hence mosquito suitability* | Minimum, maximum, annual mean, seasonality in solar radiation | [1] |
| Climate - temperature<br><br>*Impacts mosquito suitability and incubation periods* [8] | Annual minimum / maximum/ average (mean or median) land surface temperature | [1–6] |
| | Mean temperature in the wettest/ driest/ coldest/ warmest quarters | [1] |
| | Minimum temperature in the coldest month | [1] |
| | Maximum temperature in the warmest month | [1] |
| | Seasonality in temperature / annual temperature range | [1] |
| Habitats and species - Urban mosquitos<br><br>*Urban dwelling mosquitos such as* Aedes aegypti *are unlikely to drive YFV in Brazil unless there is an appreciable urban cycle* | *Aedes aegypti* habitat suitability | [7] |
| | *Aedes aegypti* temperature suitability | [7] |
| Habitats and species - sylvatic mosquitos | Maximum probability of mosquito vector occurrence (*Haemogogus leucocelaenus*, *Haemagogus janthinomys* and *Sabethes chloropterus*) | [6] |
| | Locations of vectors (*Haemagogus janthinomys* Dyar; *Haemogogus* (*Conopostegus leucocelanus* Dyar and Shannon), *Haemagogus spegazzinii* Brethes, *Haemagogus janthinomys*, *Sabethes chloropterus*, and *Sabethes cyaneus* | [4,5] |
| Habitats and species - neotropical primate | Agricultural and neotropical primate overlap (Calculated in the reference as: Proportion of total area that was both in agricultural use and within a genus range and summed across all nine considered genera, resulting in a value of 0–9 per municipality per year) | [6] |

| | | |
|---|---|---|
| *Neotropical primates are considered to be the primary amplifying host amongst which yellow fever virus transmits* | Neotropical primate species richness / distribution (*Ateles*, *Aotus*, Alou*atta*, *Saimiri*, *Cebus*, *Callicebus*, *Callithrix*, *Saguinus*, and/or *Lagothrix*) | [2–7] |
| Human | Human vaccination coverage | [2,5,9] |
| | Population density | [2,6] |
| | Population total | [2] |
| Human - impact on land use<br><br>*Could influence likelihood of human infections and/or neotropical primate or mosquito ecology* | Distance to nearest road | |
| | Deforestation | [3,5] |
| | Land use (greatest proportion biome, and/or proportion of one or more of: Evergreen broadleaf forest, urban and built up, and cropland mosaics land cover classes, "Frontier" [land use zones with potential for disruption], and/or tropical vs non-tropical) | [3–5,5,7] |
| | Net primary productivity | [1] |
| | Fire areas / fire density (as a proxy for land use change) | [2,6] |
| | Distance to the nearest road/ accessibility | [1] |
| Physical geography<br><br>*Likely affects the suitability for mosquitos and primates* | Altitude - lowest point about sea level | [3] |
| | Altitude - proportion below 2300m | |
| | Altitude - average | [1,2,4–7] |
| | Latitude | [3] |
| | Terrain slope | [1] |
| | Height above nearest drainage | [1] |
| | River basin areas | [10] |
| | Compound topographic index | [1] |
| Habitats and species - mosquito or primate<br><br>*Variables here likely affect suitability for hosts* | Average distance to forested patch | [4] |
| | Median annual EVI (enhanced vegetation index) | [2,7,11] |
| | NDVI (normalised difference vegetation index) | [6] |
| | Land cover | [2] |
| | Percentage of Ombrophilous Dense Forest | [1] |
| | Major habitat type (tropical vs sub-tropical) | [3] |
| | Percentage of deciduous forest | [1] |
| | Percentage forest cover | [1,2] |
| | Leaf area index | [1] |
| | Percentage of Ombrophilous Dense Forest | [1] |
| Additional variables not | Distance between each region (symmetrical matrix of values) - *geographically proximate* | N/A |

| | | |
|---|---|---|
| considered in cited previous studies | *locations may be more likely to spread virus due to wave-like spread* [5] | |
| | Length of border shared between contiguous regions - *longer geographic border may be more likely to result in more frequent spillovers between locations*[5] | N/A |
| | Movement of people between each region (asymmetrical matrix of values) - *human migrations may be important if urban cycle is involved* | N/A |
| | Geographic area of each region - *normalising correlate (more movements between larger areas)* | N/A |
| | Primate density of key genera - *primate density may be important in sustaining YFV transmission (with an effect that is not captured by distribution alone)* | N/A |
| | Nighttime lights / GDP per capita | N/A |
| | Proportion or absolute number of people living in rural areas or working in rural occupations - *possible proxy for exposure* | N/A |

**Supplementary Note 2: Involvement of different neotropical primate genera**

YFV surveillance in NPs is conducted through diagnostic testing of carcasses. In theory, higher mortality rates and easier detection of larger carcasses from *Alouatta spp.*[12] may lead to biased understanding that this genus is more important than other genera in YFV spread. Carcasses of NPs that live closer to urban areas, including *Callithrix spp*., may also be over-represented in detections compared to some other species[13]. Investigating genus-species transmission rates is important to understand spillover risk, because NP genera show differences in the degree of urbanisation they can tolerate (and hence their overlap with humans)[14]. We analysed sequences from NP with known host family or genus (n = 373, **Extended Data Figure 1**) to explore whether *Alouatta spp.* were more likely to contribute to onwards circulation of YFV than other NP host groups. Specifically, we used two different phylogenetic approaches to investigate the relative importance of different NP taxa as amplifier hosts.

We initially extracted from the full alignment those sequences for which information on the NP genus was available (n = 373). We grouped these sequences into *Alouatta spp.* (n = 271), *Callithrix spp*. (n = 64), and "other" (n = 38). We subsequently subsampled this dataset five times, to generate five different alignments that each contained 38 taxa from each NP group. This sub-sampling was conducted to reduce possible bias associated with a higher chance of sampling genera that are (i) more likely to die following YFV infection and therefore more likely to be detected during carcass-focused surveillance relative to true infection prevalence (*e.g., Alouatta spp)*, or (ii) relatively more capable of inhabiting urban areas (*e.g., Callithrix spp.*), and therefore more likely that carcasses are reported. Such sampling bias is known to affect discrete trait reconstruction approaches, in that groups that are sampled more relative to true infection rate are more likely to be recovered as the source of viral lineages[15].

We used both of the following approaches to analyse all 373 sequences, and then separately analysed each of the five subsampled alignments. First, we used the discrete trait reconstruction approach implemented in BEAST 1.10.5[16], using Bayesian stochastic search variable selection (BSSVS) to identify transitions required to explain the lineage movement between NP groups, and to reconstruct Markov jumps and rewards[17]. We adopted the same parameters described above and an asymmetric substitution model for host group. We used SpreaD3[18] to calculate Bayes factor support for each transition. For the full analysis of 373 sequences, we repeated this analysis whilst randomly permuting the NP group state at tips during the MCMC analysis, in order to evaluate whether higher sampling intensity of some NP groups could be excluded as causing support for particular transition rates[19]. Following[19], we used the inclusion probabilities from the trait-permuted analysis as a prior probability to calculate an adjusted Bayes factor that better accounts for sampling bias.

We next used a structured coalescent approach, as implemented in the BEAST2 v2.6.4 package, MASCOT v2.2.0 [20,21]. MASCOT was chosen instead of BASTA because whilst both MASCOT and BASTA approximate the structured coalescent, MASCOT provides a more accurate approximation[15,22]. To model sequence evolution, we used the HKY substitution model[23] with gamma-distributed rate heterogeneity[24] and an uncorrelated lognormally distributed relaxed clock model[25]. We considered the following model settings: (i) asymmetric migration rates and unequal effective population sizes across host types, (ii) asymmetric migration rates and equal effective population sizes across host types, and (iii) symmetric migration rates and equal effective population sizes across host types. We performed the analyses with these settings to check whether different constraints on the structured coalescent would affect the results. Subsequently, we investigated whether biased sampling across host types would influence the analyses. The five subsampled datasets were used to check that the pattern observed in one subsample was not purely due to sampling variability. From each subsampled dataset, we estimated the transmission history among the host genera using MASCOT with asymmetric migration rates and unequal effective population sizes across host types. The model for sequence evolution was the same as above.

We obtained conflicting results from the two different phylogenetic analyses of all NP-associated sequences. Specifically, discrete trait phylogenetic analyses of all 373 sequences suggested that *Alouatta spp.* were most likely to transmit YFV to other host groups, whereas structured coalescent approaches suggested that non-*Alouatta*, non-*Callithrix* NPs were more likely to transmit YFV. However, analyses of subsampled datasets indicates that both of those results may have been affected by sampling bias (**Extended Data Figure 10**). We therefore cannot determine whether *Alouatta spp.* are more important “amplifier hosts” of YFV than NPs that do not die as frequently, such as *Callithrix spp*. Investigating the feasibility of non-invasive methods for immune surveillance and YFV detection in well-characterised populations of live NPs would be valuable to enable fuller understanding of YFV dynamics in different NP species; for example, leveraging xeno-surveillance methods[26].

**Supplementary Note 3: Viral RNA extraction and detection procedures.**

We extracted total RNA from samples using commercially available kits. The choice of kit varied according to sample type and institution.

*Fiocruz:* We extracted RNA from tissue or sera using MagMAX Pathogen RNA/DNA kits according to the manufacturer's instructions. For tissue samples, we disrupted 30 mg tissue in 600 uL of lysis buffer, and carried 115 uL of centrifuged lysate supernatant forwards (following the manufacturer's protocol for fecal samples). For sera samples, we used 200 uL of sera. We used a KingFisher Flex extraction robot for all extractions.

*FUNED:* We extracted RNA from tissue or sera using QIAsymphony DSP Virus/Pathogen kits on a QIAsymphony extraction robot, following the manufacturer's recommendations. For serum or plasma samples, we used 250 uL of sample. For tissue samples, we cut approximately 2 mm diameter tissue sections using a disposable scalpel. We homogenized these using a 5 mm stainless steel bead in 500 uL minimum essential medium (MEM) for 15 seconds at 4.0 m/s on a FastPrep-24™ 5G. We centrifuged the lysate for 10,000 g for 2 minutes to pellet cellular material, and completed the extraction according to the manufacturer's instructions using 250 uL of supernatant.

*IAL*: We extracted RNA from 30 mg of each neotropical primate and human tissue sample using QIAamp RNA Blood Mini Kits, and from 140 uL of each serum sample using QIAamp Viral RNA Mini Kits, according to the manufacturer's instructions. A small number of samples were cultured prior to sequencing (listed on **Data S13**). For culture, we macerated tissue samples using a FastPrep-24™ 5G. We diluted supernatant in L-15 medium containing 2% FBS, penicillin (100 units/mL) and streptomycin (100 ug/mL). We innoculated 20 uL supernatant into C6/36 monolayer cell cultures, and incubated these for nine days at 28 C. We extracted RNA from these cell cultures using the methods above.

*IMT-USP:* We extracted RNA from 500uL of plasma using a NucliSENS easyMag extraction robot according to the manufacturer's instructions[27].

We tested samples for the presence of YFV RNA using previously published RT-qPCR methods.

| **Institution** | **Assay** |
|---|---|
| *Fiocruz* | We detected RNA using the methods described in[28]. |
| *FUNED* | We performed RT-qPCR using GoTaq 1-step RT-qPCR System reagents on an Applied Biosystems 7500 Real-Time PCR System machine. The PCR targets the conserved YFV 5' non-coding region using the primers and probe YFall15F, YFall103R, and YFall4, following[28]. Thermocycling conditions consisted of reverse transcription at 45 ºC for 15 min, |

| | |
|---|---|
| | denaturation at 95 °C for 2 min, followed by 45 cycles of denaturation at 95 °C for 15s, and annealing and extension at 60 °C for 1 minute. We included an assay for the RNase P gene as an endogenous positive control, following the primers and probe sequences described in[29]. |
| *IAL* | We detected RNA using two different techniques[28,30]. |
| *IMT-USP* | We detected RNA following[31], but using 14ul RNA as input as described in[27]. |

### Supplementary Note 4: Details of virus genomic sequencing

Genomic sequencing

We generated YFV genome sequences using the Oxford Nanopore Technologies MinION (n = 491 sequences), or the Illumima MiSeq (n = 7 sequences) (**Data S13**).

ONT Nanopore MinION

We conducted YFV genomic sequencing using previously published and validated protocols[11,32] that were consistent across all institutions. We generated cDNA using random hexamers. Specifically, we added 7 uL RNA to 1 uL of 50 uM random hexamers (Thermofisher). We incubated the mix at 65 °C for 5 minutes to bind the hexamers to the template before placing immediately on ice. We completed reverse transcription using the Protoscript II First Strand cDNA Synthesis kit (NEB) according to the manufacturer's instructions, with incubation conditions of 25 °C for 5 minutes, 48 °C for 15 minutes, and 80 °C for 5 minutes. We amplified YFV cDNA using the multiplex PCR described in[11]. We made 25 uL reactions (1x Q5 reaction buffer (NEB), 0.015 uM each primer, 0.25 uL Q5 Hot Start High-Fidelity DNA polymerase (NEB), 2.5uL cDNA, and nuclease-free water) and amplified on a thermocycler at 98 °C for 1 minute, followed by 32 – 40 cycles of 98 °C for 30 seconds and 65 °C for 5 minutes.

For a small subset of samples with higher PCR Cts, we performed cDNA synthesis using Superscript IV instead of Protoscript II, and used 4 ul cDNA input to the PCR reactions (these altered conditions appeared to improve amplification in testing, based on strength of fluorescence of amplicons when subjected to agarose gel electrophoresis). We performed cDNA synthesis using 1 uL of 50 uM random hexamers, 1 uL of 10mM dNTP mix, and 11uL RNA. We heated this at 70 °C for 7 minutes, before placing on ice. We added 7 uL of a mastermix containing 4 uL 5x Superscript IV reaction buffer, 1 uL of 100 mM DTT, 1 uL of RNAse OUT, and 1 uL of Superscript IV enzyme. We incubated this at 23 °C for 10 minutes, 50 °C for 45 minutes, 55 °C for 15 minutes, and 80 °C for 10 minutes to synthesise cDNA. We subsequently added 1 uL RNAse H to remove the RNA in the RNA:cDNA hybrid, and incubated at 37 °C for 20 minutes.

We purified DNA in each PCR reaction separately using 1x Ampure XP SPRI beads, eluted the DNA into 20uL of nuclease-free water, and quantified 1ul of product using the Qubit dsDNA HS assay kit on a Qubit 3.0 or a Qubit 4.0 fluorometer. We attempted to sequence PCR products only where the concentration appeared to indicate successful amplification (typically no less than 4 ng/uL, though determined flexibly depending on number of PCR cycles, tissue type, and performance of other sequenced samples with similar storage conditions).

We diluted amplicons in nuclease-free water such that 10 ng of each sample (i.e., 5 ng amplicons generated using each multiplex PCR) was carried forward into a library. In every library we included a negative control (nuclease-free water) that was processed along with the correspondent batch of samples from the point of cDNA synthesis to sequencing.

We prepared libraries using 23 samples and 1 negative control according to the 'Baseline' protocols Steps 16 - 22 in dx.doi.org/10.17504/protocols.io.bdp7i5rn[33], but with the following minor differences to reaction volumes, reagents and incubations. Specifically, we performed end repair and dA-tailing using 20uL normalized amplicons, 2.8 uL of Ultra II End Prep Reaction Buffer, and 1.2 uL of Ultra II End Prep Enzyme Mix (NEB). We barcoded dA-tailed amplicons using 24 uL of this reaction mix, and 2.5uL native barcode (ONT kits NBD104 and NBD114), 27.5 Ultra II Ligation Mix and 1 uL Ultra II Enhancer (both from NEBnext Ultra II Ligation Module). This reaction was completed at room temperature for 10 minutes, followed by 65 °C at 10 minutes to denature the ligase. We ligated the AMII adapter for 20 minutes at room temperature, using 44 uL of the pooled amplicons using 5 uL of ONT AMII adapter mix (SQK-LSK109 ONT kits), 50 uL Ultra II Ligation mix, and 1 uL of Ultra II Enhancer. All bead cleanup steps were performed at 1x ratio, and quantification was completed using the Qubit dsDNA HS assay kit.

We prepared a FLO-MIN106 flow cell according to the manufacturer's instructions, and loaded 30 ng of prepared library. We visually monitored the output of sequencing using Rampart (https://github.com/artic-network/rampart), and stopped the sequencing run after appropriate depth of coverage had been achieved (corresponding to >100x in most genomic regions of most barcodes, though regions where depth increased unusually slowly or not at all during the sequencing run were considered to have likely failed amplification during PCR and were ignored in this assessment).

We generated consensus sequences using previously published protocols developed by the ARTIC network (https://github.com/artic-network)[11,32–34]. We basecalled the raw fast5 data using Guppy version 3.0.3 (ONT). We demultiplexed reads and trimmed adaptor and barcode sequences using Porechop version 0.2.3 (https://github.com.rrwick/Porechop). We used bwa[35] to map reads to a reference genome (Genbank accession number JF912190). We identified variants for the reference genome using nanopolish version 0.11.1 variant caller[36], replacing sites with <20x coverage with an ambiguous base (N).

<u>Illumina MiSeq</u>

Although we conducted the vast majority of sequencing using the ONT MinION, we also sequenced seven samples at IMT-USP using an Illumina MiSeq according to the detailed protocol in[11]. In brief, we centrifuged samples at 20,000 g for 20 minutes, and filtered them through a 0.45 um filter to remove host cells. We treated the filtrates with DNase and RNase to degrade extracellular nucleic acid present outside of virions, and extracted RNA using a Maxwell 16 robot. We completed cDNA synthesis using 50 pmol dodecamer random primers, and AMV reverse transcriptase. We synthesized the second strand of DNA using DNA Polymerase I Large Fragment. We prepared a library using Nextera XT Sample Preparation kits and dual barcoding, and sequenced the library using an Illumina MiSeq to generate 2 x 300 bp paired end reads. We mapped demultiplexed reads to a reference genome (GenBank accession JF912190) using bwa-mem[37] and called variants using the Genome Analysis Toolkit[38] for sites with a minimum depth of 3x.

**Supplementary Note 5: Confirmation of adequacy for molecular clock phylogeographic analyses**

We checked for the presence of appropriate temporal signal for molecular clock analyses using TempEst v1.5.3[39] (**Extended Data Figure 1D**), and by confirming that TMRCAs for analyses generated using the outbreak clade sequences were similar to those generated for that clade using the full dataset of all SA1 genomes.

Phylogeographic inference can be biased by sampling[15,40]. To investigate the degree of sampling bias in our dataset, we determined whether the number of confirmed infections was a good predictor of the number of sequences obtained from each mesoregion, state or neotropical primate genus using Spearman's rank correlation coefficient as implemented in R version 4.0.5[41] using RStudio version 1.4.1106[42]. There was a significant correlation between number of sequences and number of identified confirmed infections in all analyses, indicating that our sequence selection was not highly biased relative to confirmed infections (**Extended Data Figure 8**). This does not, however, exclude that the number YFV infections confirmed by diagnostic testing may be biased relative to true infection counts.

**Supplementary Note 6: Grouping of sequences into discrete locations.**

**Mesoregion boundaries:** We grouped samples by mesoregion. Brazilian mesoregions are defined by the Brazilian Institute of Geography and Statistics (IBGE)[43] and consist of 136 groups of neighbouring municipalities with common characteristics. The sequences used here fall within 33 different mesoregions.

**K-means clustering:** We partitioned samples by k-means clustering into *k* distinct, non-overlapping groups using the R 'stats' package[41]. This was done by placing *k* sets of points randomly in space among our samples that serve as proposed centroids of groups. Each sample is grouped based on its nearest centroid, which is determined by the shortest Euclidean distance. After all samples are grouped based on a shared centroid, a new centroid is determined using the samples within each group. New groupings are then determined accordingly based on the shortest Euclidean distance between each sample and the new centroids. This process is iterated until the groupings stop changing.

**Hierarchical clustering:** We partitioned samples by agglomerative hierarchical clustering as implemented in the R 'stats' package[41]. A sample cluster is created based on the shortest Euclidean proximity between a pair of samples. This was repeated until all the samples are agglomerated into a cluster.

For both the k-means and the hierarchical clustering, we geocoded each sequence sampling location to the centroid of the sampling municipality, in order to generate clusters in which samples originating from the same municipality always appeared in the same cluster (thereby facilitating subsequent extraction of raster covariates along municipal boundaries). We visually evaluated groupings produced using values of k = 0 to k = 40, and calculated the average and maximum distance of each sample to the centroid of each group. A sharp natural breakpoint, in which distance between group centroids is unaffected by addition of further groups, was not present for either k-means or hierarchical clustering (**Extended Data Figure 9**).
We therefore chose to use k = 25 for both clustering algorithms for consistency (**Data S3**). This number was somewhat arbitrary but was chosen as a reasonable balance between having sufficient virus genomic sequences per group without overly homogenising possible environmental differences between spatial areas by grouping distantly separated municipalities.

**Supplementary Note 7: Pre-processing of rasters prior to data extraction for phylogeographic GLM**

We calculated five variables of forest loss and gain from the Global Forest Change v1.7 dataset[44]. The data available on forest loss consist of a single raster dataset where the grid cell value represents the main year in which forest loss was recorded (2001-2019). The available data on forest gain consist of a binary dataset (gain/no gain) for the time period 2000-2012. From these data, we calculated the proportion of grid cells in which there was any gain, any loss, or any churn (gain and loss) between 2000 and 2012, and also 2000 and 2017.

We calculated the summed population total and mean population density from 2016 WorldPop unconstrained gridded population[45] using a zonal statistics calculation. We estimated the population in rural areas by summing the gridded population cell values in areas classified as rural based on the GHS SMOD dataset[46].

For each spatial unit, we calculated three elevation variables: minimum elevation, maximum elevation and proportion of the spatial unit area at an elevation of below 2300m. Prior to calculating these summary statistics, we cleaned the elevation data to substitute negative values (associated with rivers and coastal areas - considered to be anomalous) with zero values. Negative values in one location associated with a deep, open cast mine were considered to not be anomalous, and left as negative elevation values.

We calculated two landcover variables from each of three landcover classes from the ESA CCI 2015 Landcover product[47]: urban (class 190), mosaic vegetation (class 40) and herbaceous (class 11). For each landcover class of interest, we created binary rasters in order to calculate the proportion of the spatial unit area of each landcover type. In addition, we calculated the distance to the edge of each landcover type, with positive values for distance outside each landcover type, and negative values associated for distances within each landcover type. From the distance to edge of landcover rasters, we calculated a mean distance value for each spatial unit.

For the time period 2015-2018, we created a mean nighttime lights brightness raster from annual gridded datasets of nighttime lights, based on harmonised data from the Visible Infrared Imaging Radiometer Suite (VIIRS) Day/Night Band (DNB)[48].

As a proxy measure for accessibility, we used a gridded dataset of distance to roads (3 arc seconds, approximately 100m spatial resolution) based on 2016 road features from OpenStreetMap (WorldPop, 2018) to calculate mean distance to roads per spatial unit.

We acquired all bioclimatic variables from WorldClim[49], providing data for the time period 1970-2000, and generated annual averages from monthly or quarterly values.

Neotropical primate genera presence variables, for specific genera occurring in the Atlantic forest (*Sapajus*, *Alouatta*, *Brachyteles*, *Callicebus*, *Callithrix*, *Leontopithecus*) were provided as a gridded dataset with an estimated (modelled) count of primates per grid cell at a spatial resolution of approximately 1km[50]. Data on species richness at the municipality level were available as a CSV file at the municipality level with the mean number of species and the maximum number of species found[802].

For mining data, we considered clusters of municipalities in which any municipality had a substantial contribution to municipal gross domestic product (GDP) to have the “presence of mining industry”. We defined a “substantial” contribution to be where was mining the primary, secondary or tertiary contributor to GDP in at least 2 years between 2014-2016. For vaccination coverage, estimates of the number of people in each cluster, and proportion of vaccination coverage, were calculated for the cluster as a whole using information on the number of people in each municipality.

**Supplementary Note 8: Extraction of data for skygrid-GLM.**

We obtained measures of vegetation greenness from the Moderate Resolution Imaging Spectroradiometer (MODIS) imagery supplied by NASA[51]. The MOD13c2 version 6 hdf format product provides monthly global images of several vegetation indices at approximately 5km resolution, from which the Enhanced Vegetation Index (band 1) was extracted. We calculated mean monthly values of this metric for each of the land areas required in Python using ESRI ArcGis v10.8 functions ExtractSubdatset and ZonalStatisticsasTable. We obtained sea surface temperature (SST) anomalies from NOAA (https://www.cpc.ncep.noaa.gov/data/indices/sstoi.indices). Anomalies are calculated by subtracting the month-specific mean SST for the period 1981-2010 to each SST value for the Niño 3.4 region (5°North-5°South; 170-120°West). We retrieved all other climatic data in netcdf files at monthly time steps from the fifth generation ECMWF atmospheric reanalysis of the global climate (ERA5)[52] through the Copernicus Climate Data Store (https://cds.climate.copernicus.eu/) at a 0.25 x 0.25 arc degree resolution for the whole globe.

**Supplementary Note 9: Hypothesis testing based on the continuous phylogeographic reconstruction**

For the hypothesis testing analyses based on the continuous phylogeographic reconstruction, a discipline also coined as landscape phylogeography[53], we used three different approaches[54–57] all implemented in the R package "seraphim"[58,59]. All these approaches require use of a null dispersal model to which the inferred dispersal history of YFV lineages can be compared. In brief, we sample 1,000 trees from the posterior distribution, and simulate a forwards-in-time relaxed random walk (RRW) process from the inferred tree root location using sampled precision matrix parameters estimated during continuous phylogeography. Tree topologies and root location are therefore identical in each pair of trees from the inferred and simulated tree sets, and only the geographic locations of internal and tip locations differ. We constrain the simulation such that each simulated internal node location remains on land. We used the "simulatorRRW1" function of the R package "seraphim" for all simulations[58]. For all analyses below, we used the sampled set of 1,000 trees ("inferred trees") and compared information from each tree to the corresponding tree in which node locations had been simulated ("simulated tree").

We use first the approach of Dellicour et al[54] to determine whether YFV tends to preferentially circulate within the same ecoregion, or whether dispersal is unaffected by ecoregion. We estimated the number of ecoregion switching events for each inferred tree, based on the ecoregions associated with the node location at the two nodes that define each branch. We tested whether the number of switching events was substantially greater or lesser than that in each corresponding simulated tree using a previously described approximation of Bayes factor support[55,60]: $BF = (p/(1-p))/(0.5/(1-0.5))$, where $p$ is the frequency at which the estimated number of switches from the inferred trees is greater that the estimated number of switched in the simulated trees.

For the second analysis, we investigated whether YFV lineages tended to preferentially circulate within or avoid areas associated with particular environmental characteristics. We calculated the average value of each environmental variable across all phylogenetic nodes of an inferred tree. Average values from each inferred tree therefore defined a posterior distribution of environmental values associated with dispersal. We formally compared values obtained through inferred trees and their corresponding simulated trees using the above formula for Bayes factor support[55,60]. To test if a specific environmental factor $e$ tended to attract or repulse viral lineages, $p_e$ was defined as the frequency at which the environmental values from inferred trees were greater than values from simulated trees, whereas the reverse was used to test if the environmental factor impedes lineages.

Finally, we considered whether YFV dispersal was made faster ("conducted") or made slower ("resisted") by each environmental variable in **Data S14**. We use the approach described in Dellicour et al[57]. In brief, for each branch in the inferred and simulated trees we calculate an environmentally scaled distance using the Circuitscape algorithm[61], a path model that accommodates uncertainty in the travel route. An environmentally scaled distance is calculated first from the raster of the environmental variable, and second from a uniform "null" raster whose cell values are all set to "1" (rasters are publicly available as indicated in the main text **Data Availability** statement). The environmentally scaled distance is a spatial distance that is weighted according to the values of the underlying environmental raster. Therefore, environmentally scaled distances computed on the null raster constitute a proxy for geographical distances. Each environmental variable must be considered twice: once as a conductance factor that facilitates movement, and once as a resistance factor that impedes it.

A statistic $Q$ is defined as the difference between the coefficient of determinations obtained when branch durations are regressed against the environmentally scaled distance calculated against the (i) informative raster, and (ii) the null raster. Distributions of $Q$ values are calculated for the inferred and simulated tree sets. Environmental variables were considered potentially explanatory where greater than 90% of $Q$ values were >0 and where regression coefficients of regressions against null and informative rasters were positive. Support for the importance of each potentially explanatory environmental variable to facilitate or impede dispersal velocity is estimated using an approximation of Bayes factor support as described above[55,60]. To assess the strength of conductance or resistance for each variable, we repeated each test using rasters with cell values that had been subjected to rescaling by a factor $k$[55,57], with the transformed value $v_t=1+kv_0$, where $v_0$ is the original value. We used three different values of $k$: 10, 100 and 1,000.

### Extended Data Figures

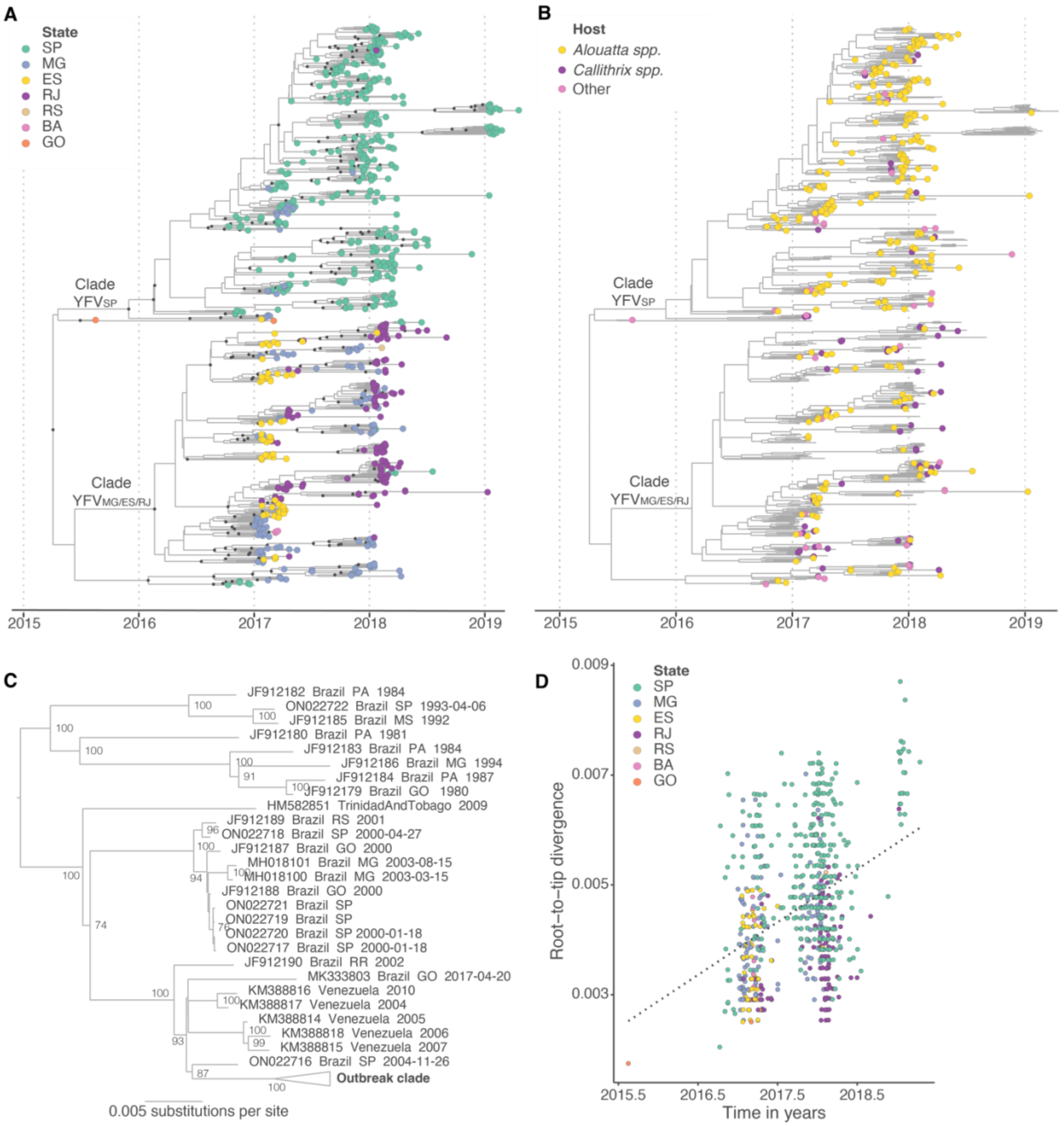


**Extended Data Figure 1. A and B:** Maximum clade credibility (MCC) tree of Brazilian outbreak clade. **A**: Colours at tips denote geographic federal state of sampling (SP: São Paulo, MG: Minas Gerais, ES: Espírito Santo, RJ: Rio de Janeiro, RS: Rio Grande do Sul, BA: Bahia, GO: Goiás.) Internal nodes with posterior support >0.8 are shown in grey. **B:** Colours at tips denote host group used in discrete trait analyses (yellow: *Alouatta spp.*, purple: *Callithrix spp.*, pink: other neotropical primate species). No tip shape is shown for human or mosquito samples. **C:** Maximum likelihood phylogeny of all curated YFV SA1 sequences. The monophyletic outbreak clade is marked in bold. Bootstrap scores >70 are shown. Only one sequence sampled since 2010 in Brazil falls outside this monophyletic clade (MK333803). As previously observed, this sequence instead forms part of a closely related clade containing sequencing sampled in Venezuela[10]. Whilst genetically distinct from the outlier clade, it is not an outlier in root-to-tip divergence against time analyses. However, MK333803 was sampled in a municipality in northwestern Goiás that borders Mato Grosso state and is therefore geographically isolated from all other samples considered here, so was excluded on this basis from most analyses. **D:** Root-to-tip divergence against time of sequences in the Brazilian outbreak clade. The regression obtained from TempEst [39] is shown.

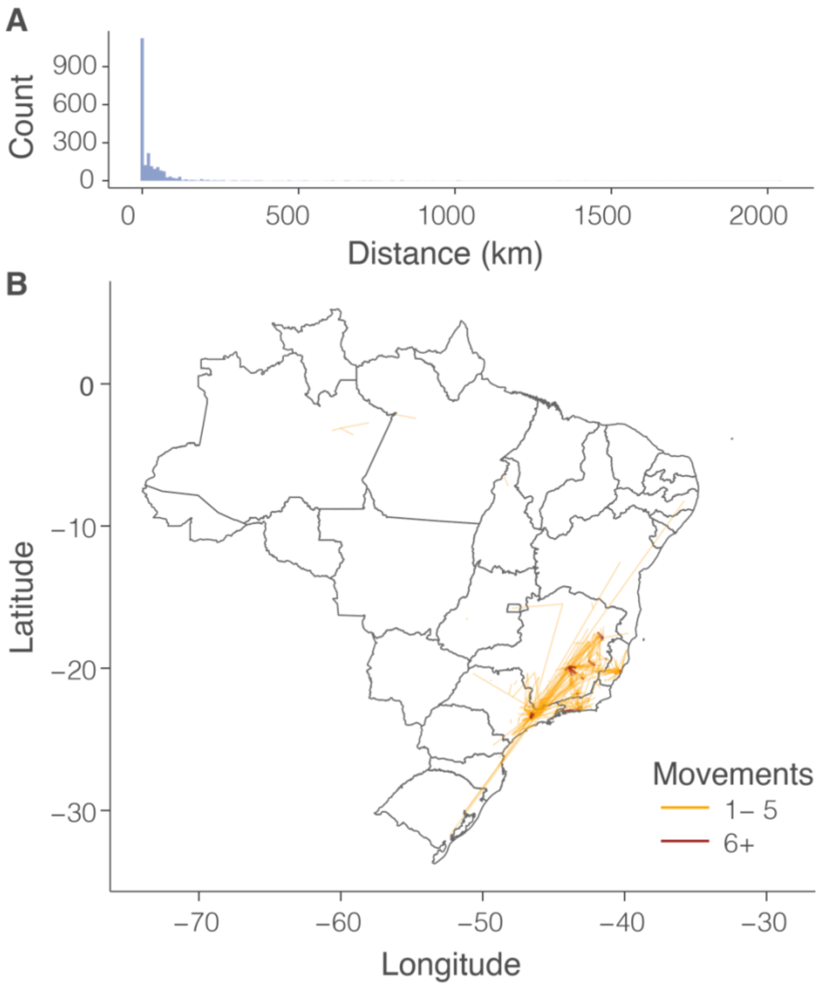


**Extended Data Figure 2. Reported movement of patients with confirmed YFV infection. A:** histogram of the movement distance per patient from the notification location to the presumed infection location. **B:** number of movement events between every pair of municipalities. Trajectories are marked in red when over five journeys are reported.

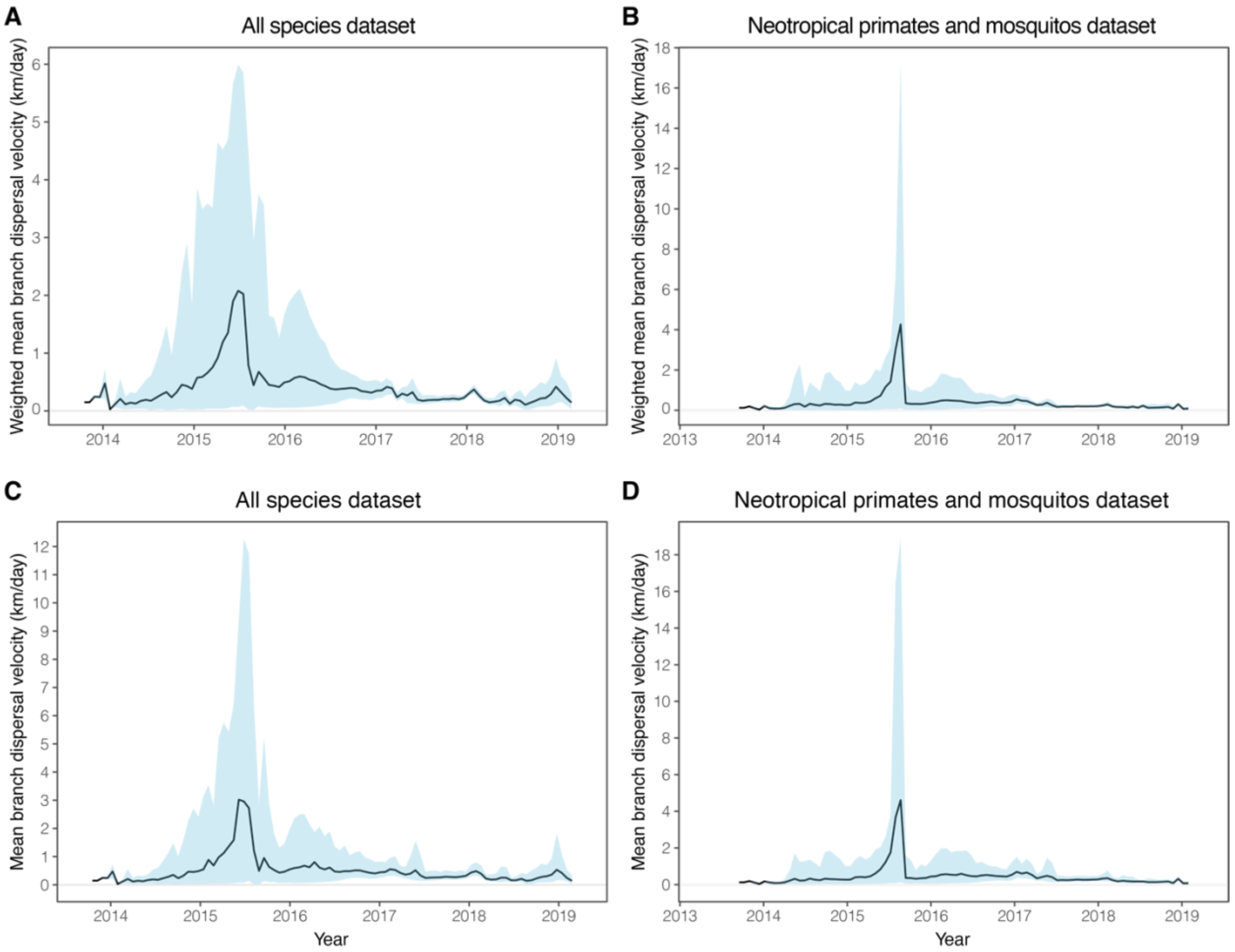


**Extended Data Figure 3. Estimated lineage dispersal velocity.** Black lines show mean estimates, and blue ribbons show 95% HPD intervals. **A:** weighted lineage dispersal velocity for full dataset (humans, mosquitos and NP samples). **B:** mean lineage dispersal velocity for full dataset (humans, mosquitos and NP samples). **C:** weighted lineage dispersal velocity for non-human dataset (mosquitos and NP samples). **D:** mean lineage dispersal velocity for non-human dataset (mosquitos and NP samples).

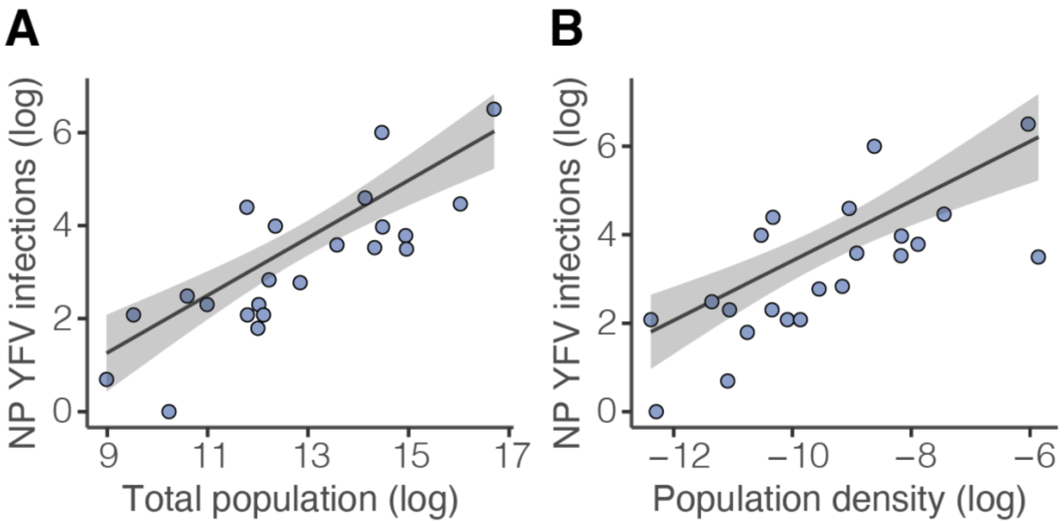


**Extended Data Figure 4. Correlation between number of YFV infections confirmed in NPs and two measures of human population presence. A:** total human population. **B:** human population density. Values shown here are extracted from municipalities grouped through hierarchical clustering (described in **Supplementary Materials and Methods**, section 'Clustering of sequences into discrete groups'). The regression was fit using a negative binomial GLM with a log link, and 95% confidence intervals are indicated with a ribbon. Both relationships are significant at $P < 0.001$.

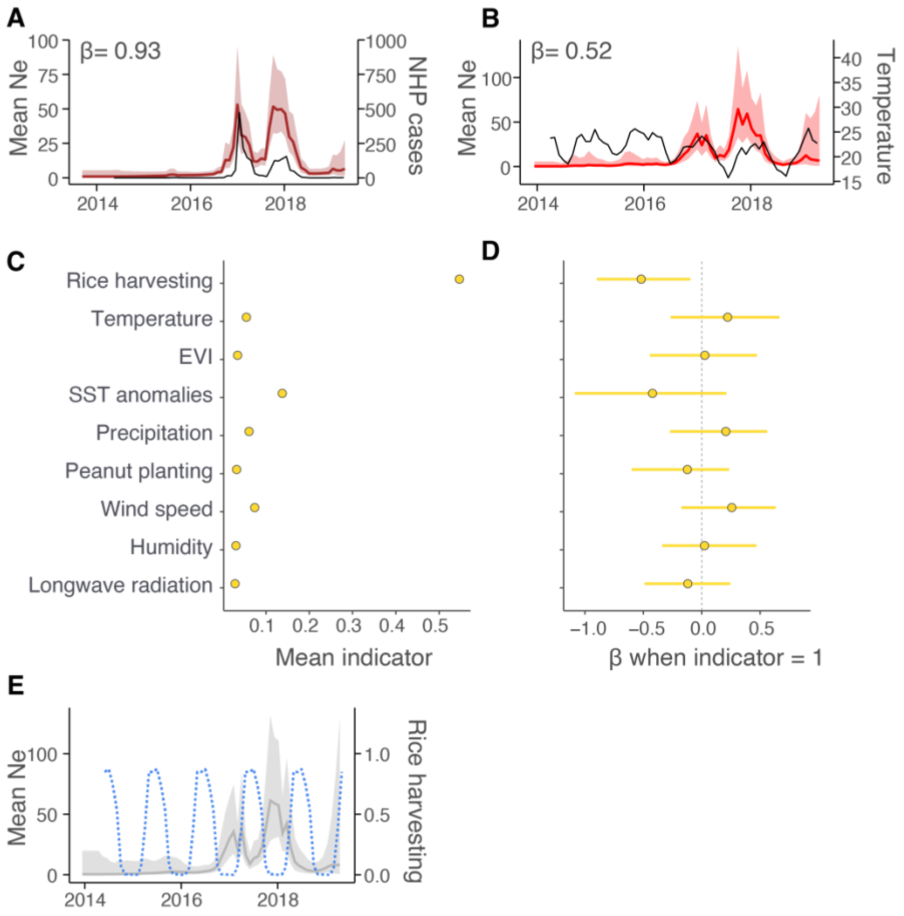


**Extended Data Figure 5. Factors associated with YFV effective population size. A** and **B:** Associations between mean viral effective population size ($N_e$) and **A:** number of confirmed YFV infections in neotropical primates or **B:** mean annual temperature (°C). For both **A** and **B**, coloured lines show the mean $N_e$ estimated from viral sequence and covariate data (left hand axis), and ribbons show the 95% HPD. Black lines show the values of covariates at each time point. Covariate values were extracted per month for the region in which transmission was occurring based on the results of continuous phylogeographic analysis. **C, D and E:** results of skygrid-GLM with one-month lag (i.e., covariates from any month are used to predict virus effective population size in the subsequent month). **C** shows the mean of the indicators for each variable, and **D** shows the regression coefficient when the indicator for that variable is 1. EVI: enhanced vegetation index. **E**: Proportion of farms harvesting rice (blue dotted lines), and mean virus effective population size ($N_e$). Grey lines show mean $N_e$ estimated from viral sequence data during an analysis uninformed by covariate priors, and ribbons show the 95% HPD. Covariate values were extracted per month for the region containing the federal states of SP, ES, RJ and MG.

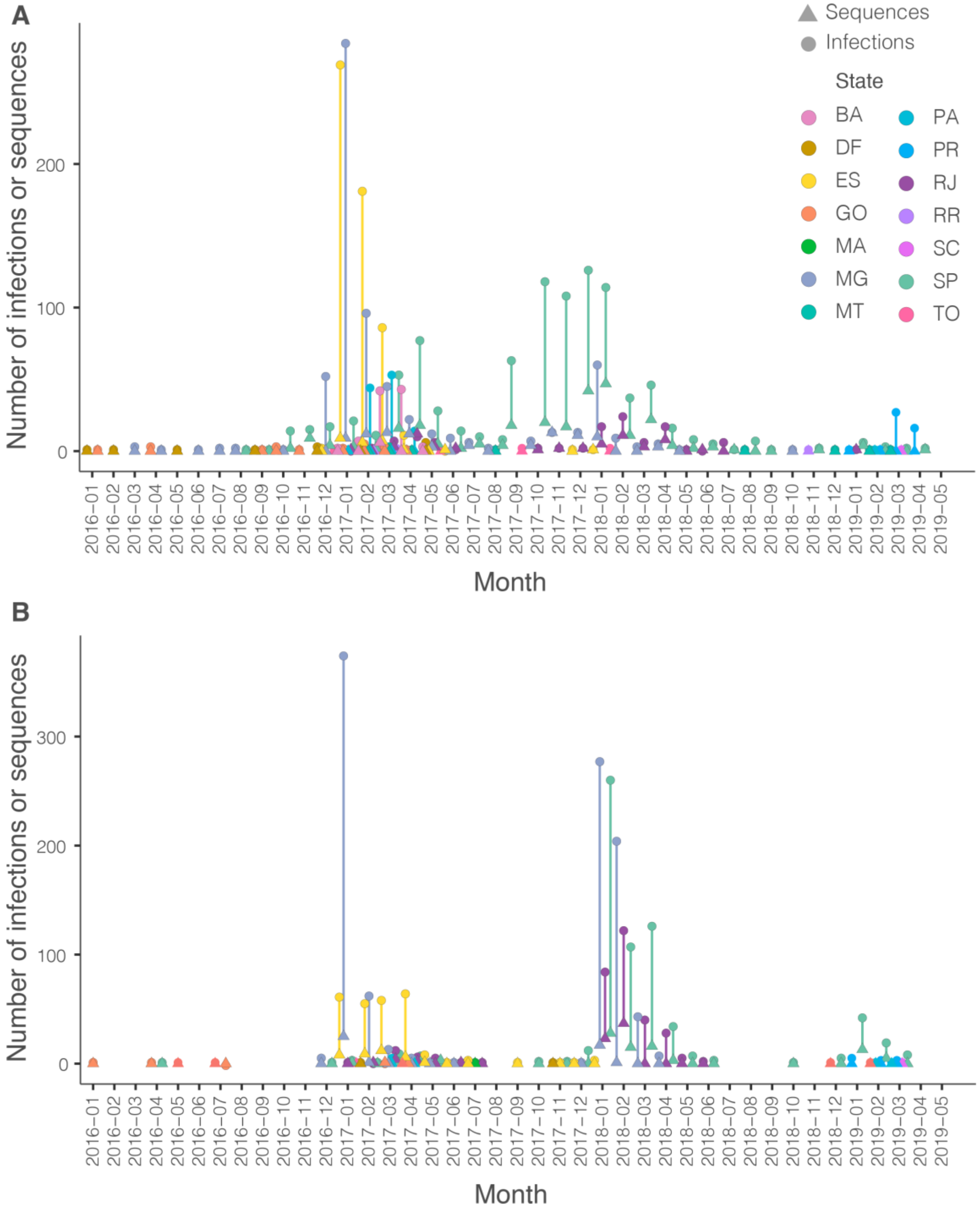


**Extended Data Figure 6. Number of confirmed infections and available sequences per state and month (Jan 2016 to April 2019).** Triangles represent the number of sequences and circles represent the number of infections. Colours indicate states. **A**: neotropical primates. **B**: humans.

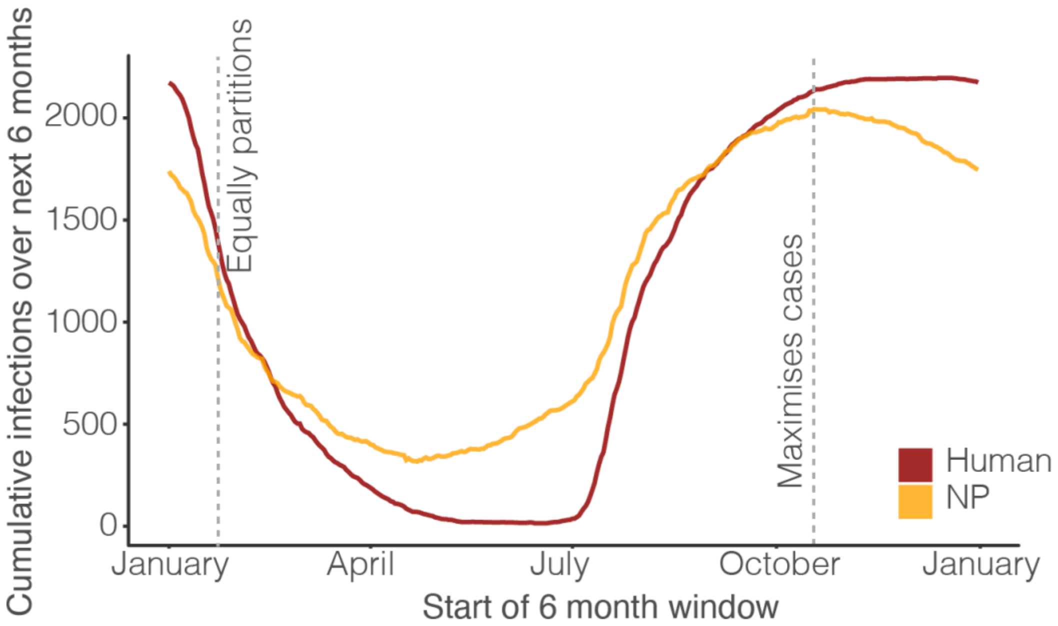


**Extended Data Figure 7.** Defining epochs used in time-varying rate analyses. Cumulative numbers of confirmed YFV infections observed in 6-month sliding windows for humans and NPs. Vertical dashed lines indicate the time of the year where placement of a transition point (i) maximises neotropical primate (NP) infections within epoch 1 and minimizes infections within epoch 2, and (ii) renders infections equally distributed across both epochs. Red = human, yellow = NP.

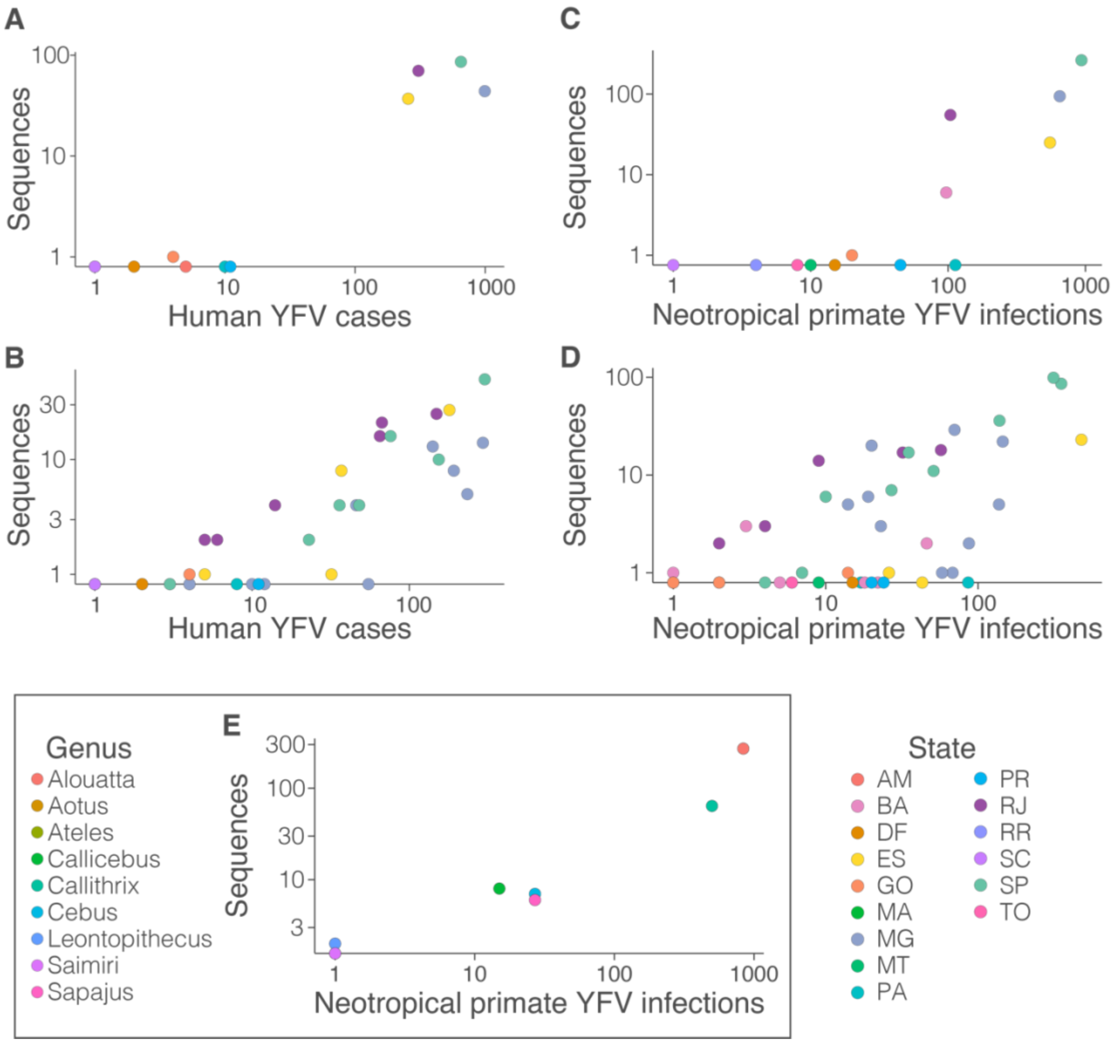


**Extended Data Figure 8. Correlation between confirmed YFV infections and number of genomic sequences. A:** per state for humans. **B**: per mesoregion for humans. **C**: per state for neotropical primates. **D**: per mesoregion for neotropical primates. **E**: per genus for neotropical primates. Points are coloured by genus (**E**) or state (**A-D**) (AM: Amazonas, BA: Bahia, DF: Distrito Federal, ES: Espírito Santo, GO: Goiás, MA: Maranhão, MG: Minas Gerais, MT: Mato Grosso, PA: Pará, PR: Paraná, RJ: Rio de Janeiro, SC: Santa Catarina, SP: São Paulo, TO: Tocantins)). All correlations are significant by Spearman's rank correlation coefficient ($p<0.005$).

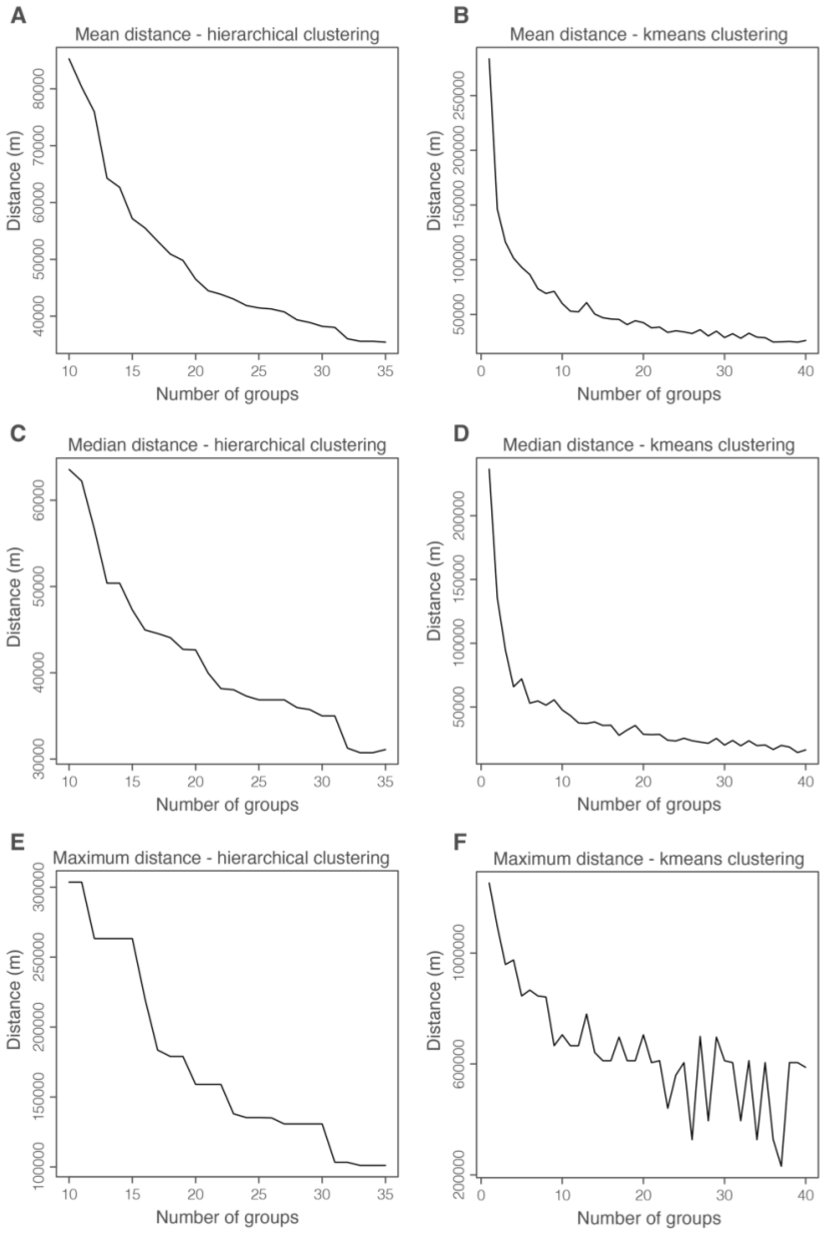


**Extended Data Figure 9. Impact of changing number of groups on distances between geocoded samples and group centroid. Panel A, C, E;** mean, median and maximum distances for groups defined using the described hierarchical clustering algorithm. **Panel B, D, F;** mean, median and maximum distances for groups defined using the described k-means clustering algorithm.

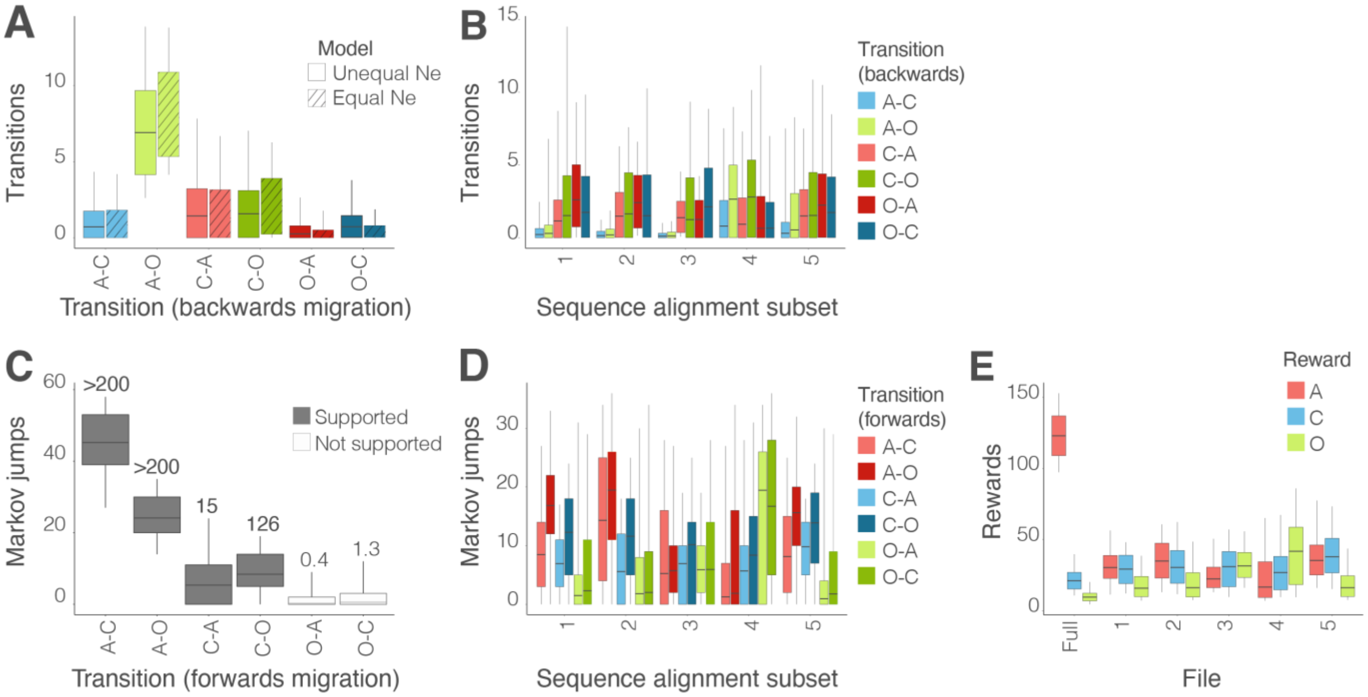


**Extended Data Figure 10. Phylogenetically reconstructed transmission between neotropical primate host groups.** **A** and **B** show results obtained from the MASCOT structured coalescent approach implemented in BEAST2, whereas **C**, **D** and **E** show results from a discrete trait reconstruction implemented in BEAST. In all figure panels, *Alouatta* is marked with 'A', *Callithrix* with 'C', and *Other* with 'O', such that transitions from, e.g., *Alouatta* to *Callithrix* are marked as 'A-C'. **A:** estimated transitions between different neotropical primate groups for models based on asymmetric rates of virus lineage movement and equal (plain) and unequal (diagonal lines) effective population sizes. **B**: transitions between different host pairs estimated using 5 different subsets of sequence alignments. **C:** estimated number of transitions (Markov jumps) associated with each pair of neotropical primate groups. Grey bars show transitions that are supported by adjusted Bayes factors, and white bars show transitions that are not supported. Bayes factors are provided. **D**: Number of transitions (Markov jumps) estimated between different host pairs estimated using 5 different subsets of sequence alignments. **E**: Estimated time spent in each state (Markov rewards) based on the full dataset, and 5 different subsets of sequences. Note that in **A** and **B**, backwards migration is shown (from tips to root of the tree), whereas in **C** and **D**, forwards migration is shown (from root to tips). Colours are used in panels **A**, **C** and **D** to show equivalency of transmission routes (e.g., backwards migration from *Alouatta* to *Callithrix* is comparable to forwards migration from *Callithrix* to *Alouatta*).

**Extended Data Table 1: Lineage dispersal velocity statistics**

| Sequence set | Mean lineage dispersal velocity, km/day (95% CI) | Weighted lineage dispersal velocity, km/day (95% CI) |
|---|---|---|
| All species | 1.23 (1.01 – 1.86) | 0.59 (0.53 – 0.66) |
| Non-human species only | 1.21 (0.97 – 1.93) | 0.60 (0.54 – 0.67) |

### Legends of Supplementary Data Files

**Data S1. (separate file)**
**Covariates chosen for phylogeographic GLMs for all analyses.**

**Data S2. (separate file)**
**Results of phylogeographic GLM analyses.**

**Data S3. (separate file)**
**Groupings of municipalities used in k-means and hierarchical clustering analyses.** Note that possible maximum value of estimated BFs is artificially limited here by the number of samples in the posterior distribution. We therefore report Bayes factors as >999 where relevant in the main text, though the true BF will often be much higher.

**Data S4. (separate file)**
**Impact of several environmental factors on the dispersal location of YFV lineages in Brazil based on the entire YFV genomic data set.** We report Bayes factor (BF) support for the association between environmental values and tree node locations. The results are based on 100 posterior trees obtained by spatially-explicit phylogeographic inference. Following Kass & Raftery (1995), we consider a BF value >=20 as strong support for a significant correlation between the environmental distances and dispersal durations (in bold), and a BF value >=3 and <20 as positive (underlined). "ENM" refers to ecological niche modelling to predict species' distributions.

**Data S5. (separate file)**
**Impact of several environmental factors on the dispersal location of YFV lineages in Brazil based on the non-human YFV genomic data set.** We report Bayes factor (BF) support for the association between environmental values and tree node locations. The results are based on 100 posterior trees obtained by spatially-explicit phylogeographic inference. Following Kass & Raftery (1995), we consider a BF value >20 as strong support for a significant correlation between the environmental distances and dispersal durations (in bold), and a BF value >3 and <20 as positive (underlined). "ENM" refers to ecological niche modelling.

**Data S6. (separate file).**
**Collinearity of covariates considered for continuous phylogeographic analyses.**

**Data S7. (separate file)**

**Collinearity of covariates considered for phylogeographic GLM, hierarchical clustering dataset.**

**Data S8. (separate file)**
**Collinearity of covariates considered for phylogeographic GLM, k-means clustering dataset.**

**Data S9. (separate file)**

**Collinearity of covariates considered for phylogeographic GLM, mesoregion dataset.**

**Data S10. (separate file)**
**Impact of several environmental factors on the dispersal velocity of YFV lineages in Brazil, based on the entire YFV genomic data set and with environmental distances computed with the Circuitscape path model.** The results are based on 100 posterior trees obtained by spatially-explicit phylogeographic inference. “C” and “R” indicate if the considered environmental raster was considered as a conductance (“C”) or resistance factor (“R”), and *k* is the rescaling parameter used to transform the initial raster (see the text for further details). For regression coefficients and *Q* values we report both the median estimate and the 95% HPD interval. The Bayes factor (BF) supports are only reported when p(*Q* > 0) is at least 90%. Following Kass & Raftery (1995), we consider a BF value >20 as strong support for a significant correlation between the environmental distances and dispersal durations (in bold), and a BF value >3 and <20 as positive (underlined).

**Data S11. (separate file)**
**Impact of several environmental factors on the dispersal velocity of YFV lineages in Brazil, based on the non-human YFV genomic data set and with environmental distances computed with the Circuitscape path model.** The results are based on 100 posterior trees obtained by spatially-explicit phylogeographic inference. “C” and “R” indicate if the considered environmental raster was considered as a conductance (“C”) or resistance factor (“R”), and *k* is the rescaling parameter used to transform the initial raster (see the text for further details). For regression coefficients and *Q* values we report both the median estimate and the 95% HPD interval. The Bayes factor (BF) supports are only reported when p(*Q* > 0) is at least 90%. Following Kass & Raftery (1995), we consider a BF value >20 as strong support for a significant correlation between the environmental distances and dispersal durations (in bold), and a BF value >3 and <20 as positive (underlined).

**Data S12. (separate file)**
**Evolutionary rates measured in epoch models.** Columns show the differences between rates for epochs 1 and 2 and 95% HPD intervals, and the median rates.

**Data S13. (separate file)**
GenBank accession numbers and available metadata of all sequences generated or used in this study.

**Data S14. (separate file)**
**Covariates considered or used for inclusion in phylogeographic GLMs and the sources of data.**

**Data S15. (separate file)**
**Raw data considered for inclusion in phylogeographic GLMs.** Where origin and destination matrices are supplied separately, values are given as from “*row name*” to “*column name*”, otherwise values are symmetrical (e.g., distance between two regions).

**Data S16. (separate file)**
**Covariates used in skygrid-GLM analyses.** 1 = included, 0 = excluded.

**Data S17. (separate file)**
Collinearity of covariates considered for skygrid-GLM, states regions.

**Data S18. (separate file)**
Collinearity of covariates considered for skygrid-GLM, regions informed by polygons obtained in continuous phylogeographic analyses.

**Data S19. (separate file)**
XML files for all phylodynamic analyses.
