## Supplementary Data (Data S1 - S19) for "Climate and land-use shape the spread of zoonotic yellow fever virus": Data S4.docx

Data S4. Impact of several environmental factors on the dispersal location of YFV lineages in Brazil based on the entire YFV genomic data set. We report Bayes factor (BF) supports for the association between environmental values and tree node locations. The results are based on 100 posterior trees obtained by spatially-explicit phylogeographic inference. Following Kass & Raftery (1995), we consider a BF value >=20 as strong support (in bold), and a BF value >=3 and <20 as positive (underlined). “ENM” refers to ecological niche modelling to predict species’ distributions.

| Environmental factor | Tendency of viral lineages to avoid circulating within specific environmental conditions | Tendency of viral lineages to preferentially circulate within specific environmental conditions |
| --- | --- | --- |
| *Aedes aegypti* (ENM) | **46.6** | 0.0 |
| *Aedes albopictus* (ENM) | 4.9 | 0.2 |
| Annual mean temperature | 19.0 | 0.1 |
| Annual precipitation | 0.3 | 3.0 |
| Aridity | 0.0 | **33.5** |
| Distance to main roads | **>99** | 0.0 |
| Elevation | 0.7 | 1.5 |
| Enhanced vegetation index (2005-18) | 0.1 | 6.8 |
| Forest loss (2000-17) | 1.1 | 0.9 |
| Land cover - herbaceous | **89.9** | 0.0 |
| Land cover - mosaic natural vegetation | 0.0 | **21.7** |
| Land cover - urban areas | 0.0 | **>99** |
| Nightlights (2018) | 0.0 | **>99** |
| Human population density | 0.0 | **>99** |
| Expected number of primates of six genera | 0.0 | **>99** |
