## Supplementary Data (Data S1 - S19) for "Climate and land-use shape the spread of zoonotic yellow fever virus": Data S10.docx

Data S10. Impact of several environmental factors on the dispersal velocity of YFV lineages in Brazil, based on the entire YFV genomic data set and with environmental distances computed with the Circuitscape path model. The results are based on 100 posterior trees obtained by spatially-explicit phylogeographic inference. “C” and “R” indicate if the considered environmental raster was considered as a conductance (“C”) or resistance factor (“R”), and *k* is the rescaling parameter used to transform the initial raster (see the text for further details). For regression coefficients and *Q* values we report both the median estimate and the 95% HPD interval. The Bayes factor (BF) supports are only reported when p(*Q* > 0) is at least 90%. Following Kass & Raftery (1995), we consider a BF value >20 as strong support for a significant correlation between the environmental distances and dispersal durations (in bold), and a BF value >3 and <20 as positive (underlined).

| Environmental factor | *k* | Regression coefficient | *Q* statistic | p(*Q* > 0) | BF |
| --- | --- | --- | --- | --- | --- |
| *Aedes aegypti* (ENM, C) | 10 | 0.057 [0.041, 0.073] | -0.026 [-0.043, -0.010] | 0.000 | - |
|  | 100 | 0.049 [0.033, 0.063] | -0.033 [-0.053, -0.015] | 0.000 | - |
|  | 1000 | 0.047 [0.033, 0.062] | -0.035 [-0.054, -0.016] | 0.000 | - |
| *Aedes aegypti* ( ENM, R) | 10 | 0.062 [0.043, 0.089] | -0.019 [-0.029, -0.008] | 0.001 | - |
|  | 100 | 0.058 [0.039, 0.084] | -0.023 [-0.035, -0.011] | 0.001 | - |
|  | 1000 | 0.057 [0.038, 0.084] | -0.024 [-0.036, -0.012] | 0.001 | - |
| *Aedes albopictus* (C) | 10 | 0.073 [0.055, 0.096] | -0.009 [-0.019, -0.001] | 0.002 | - |
|  | 100 | 0.069 [0.053, 0.093] | -0.011 [-0.024, -0.002] | 0.001 | - |
|  | 1000 | 0.069 [0.053, 0.092] | -0.011 [-0.024, -0.002] | 0.001 | - |
| *Aedes albopictus* (R) | 10 | 0.076 [0.059, 0.098] | -0.005 [-0.012, 0.002] | 0.012 | - |
|  | 100 | 0.075 [0.058, 0.096] | -0.006 [-0.014, 0.002] | 0.008 | - |
|  | 1000 | 0.074 [0.058, 0.096] | -0.006 [-0.014, 0.002] | 0.007 | - |
| Annual mean temperature (C) | 10 | 0.089 [0.073, 0.108] | 0.009 [-0.006, 0.021] | 0.091 | 8.1 |
|  | 100 | 0.089 [0.073, 0.108] | 0.008 [-0.006, 0.021] | 0.088 | - |
|  | 1000 | 0.089 [0.073, 0.108] | 0.008 [-0.006, 0.002] | 0.088 | - |
| Annual mean temperature (R) | 10 | 0.085 [0.068, 0.108] | 0.004 [-0.012, 0.023] | 0.082 | - |
|  | 100 | 0.085 [0.067, 0.108] | 0.003 [-0.012, 0.023] | 0.079 | - |
|  | 1000 | 0.084 [0.067, 0.108] | 0.003 [-0.012, 0.023] | 0.078 | - |
| Annual precipitation (C) | 10 | 0.083 [0.065, 0.104] | 0.002 [-0.013, 0.019] | 0.062 | - |
|  | 100 | 0.080 [0.062, 0.101] | 0.000 [-0.016, 0.017] | 0.046 | - |
|  | 1000 | 0.080 [0.061, 0.101] | -0.001 [-0.016, 0.017] | 0.045 | - |
| Annual precipitation (R) | 10 | 0.087 [0.070, 0.107] | 0.006 [-0.009, 0.017] | 0.086 | - |
|  | 100 | 0.086 [0.069, 0.106] | 0.005 [-0.010, 0.016] | 0.077 | - |
|  | 1000 | 0.085 [0.068, 0.106] | 0.005 [-0.010, 0.015] | 0.076 | - |
| Aridity (C) | 10 | 0.073 [0.054, 0.105] | -0.009 [-0.022, 0.019] | 0.022 | - |
|  | 100 | 0.069 [0.049, 0.102] | -0.012 [-0.028, 0.016] | 0.014 | - |
|  | 1000 | 0.069 [0.049, 0.102] | -0.013 [-0.028, 0.015] | 0.014 | - |
| Aridity (R) | 10 | 0.077 [0.049, 0.098] | -0.004 [-0.023, 0.011] | 0.031 | - |
|  | 100 | 0.075 [0.045, 0.095] | -0.006 [-0.027, 0.008] | 0.019 | - |
|  | 1000 | 0.074 [0.045, 0.094] | -0.006 [-0.027, 0.008] | 0.018 | - |
| Distance to main roads (C) | 10 | 0.075 [0.050, 0.096] | -0.007 [-0.028, 0.011] | 0.018 | - |
|  | 100 | 0.019 [0.008, 0.029] | -0.062 [-0.081, -0.043] | 0.000 | - |
|  | 1000 | 0.001 [0.000, 0.005] | -0.080 [-0.105, -0.064] | 0.000 | - |
| Distance to main roads (R) | **10** | **0.098 [0.077, 0.138]** | **0.016 [0.000, 0.046]** | **0.097** | **>99** |
|  | 100 | 0.071 [0.051, 0.103] | -0.011 [-0.031, 0.016] | 0.018 | - |
|  | 1000 | 0.056 [0.036, 0.081] | -0.027 [-0.048, 0.000] | 0.001 | - |
| Elevation (C) | 10 | 0.033 [0.021, 0.055] | -0.048 [-0.068, -0.024] | 0.000 | - |
|  | 100 | 0.009 [0.004, 0.018] | -0.071 [-0.093, -0.054] | 0.000 | - |
|  | 1000 | 0.007 [0.002, 0.015] | -0.073 [-0.095, -0.056] | 0.000 | - |
| Elevation (R) | **10** | **0.106 [0.086, 0.130]** | **0.026 [0.003, 0.047]** | **0.100** | **>99** |
|  | 100 | 0.090 [0.069, 0.116] | 0.010 [-0.014, 0.032] | 0.082 | - |
|  | 1000 | 0.088 [0.067, 0.113] | 0.007 [-0.017, 0.029] | 0.076 | - |
| Enhanced vegetation index | 10 | 0.058 [0.041, 0.078] | -0.024 [-0.036, -0.006] | 0.001 | - |
| (2005-2018, C) | 100 | 0.049 [0.035, 0.067] | -0.031 [-0.045, -0.015] | 0.000 | - |
|  | 1000 | 0.048 [0.034, 0.066] | -0.032 [-0.047, -0.017] | 0.000 | - |
| Enhanced vegetation index | 10 | 0.083 [0.060, 0.111] | 0.002 [-0.015, 0.022] | 0.061 | - |
| (2005-2018, R) | 100 | 0.082 [0.058, 0.111] | 0.002 [-0.016, 0.022] | 0.057 | - |
|  | 1000 | 0.082 [0.058, 0.111] | 0.002 [-0.016, 0.022] | 0.055 | - |
| Forest loss (2000-2019, C) | 10 | 0.082 [0.061, 0.103] | 0.000 [-0.016, 0.019] | 0.050 | - |
|  | 100 | 0.032 [0.016, 0.048] | -0.048 [-0.072, -0.030] | 0.000 | - |
|  | 1000 | 0.006 [0.001, 0.016] | -0.074 [-0.098, -0.057] | 0.000 | - |
| Forest loss (2000-2019, R) | 10 | 0.078 [0.060, 0.104] | -0.003 [-0.023, 0.024] | 0.040 | - |
|  | 100 | 0.030 [0.017, 0.047] | -0.051 [-0.074, -0.028] | 0.000 | - |
|  | 1000 | 0.021 [0.010, 0.034] | -0.060 [-0.083, -0.038] | 0.000 | - |
| Land cover - herbaceous (C) | 10 | 0.020 [0.010, 0.030] | -0.061 [-0.084, -0.045] | 0.000 | - |
|  | 100 | 0.002 [0.000, 0.008] | -0.079 [-0.100, -0.061] | 0.000 | - |
|  | 1000 | 0.000 [0.000, 0.002] | -0.081 [-0.106, -0.063] | 0.000 | - |
| Land cover - herbaceous (R) | 10 | 0.068 [0.047, 0.094] | -0.014 [-0.039, 0.012] | 0.017 | - |
|  | 100 | 0.047 [0.030, 0.069] | -0.035 [-0.061, -0.011] | 0.000 | - |
|  | 1000 | 0.043 [0.027, 0.065] | -0.038 [-0.064, -0.016] | 0.000 | - |
| Land cover - mosaic | 10 | 0.061 [0.046, 0.082] | -0.018 [-0.041, 0.001] | 0.005 | - |
| natural vegetation (C) | 100 | 0.030 [0.018, 0.048] | -0.049 [-0.076, -0.028] | 0.000 | - |
|  | 1000 | 0.009 [0.002, 0.028] | -0.071 [-0.095, -0.048] | 0.000 | - |
| Land cover - mosaic | 10 | 0.033 [0.020, 0.048] | -0.047 [-0.070, -0.031] | 0.000 | - |
| natural vegetation (R) | 100 | 0.021 [0.011, 0.034] | -0.058 [-0.081, -0.043] | 0.000 | - |
|  | 1000 | 0.019 [0.010, 0.032] | -0.060 [-0.083, -0.045] | 0.000 | - |
| Land cover - urban areas (C) | 10 | 0.097 [0.074, 0.129] | 0.016 [-0.010, 0.038] | 0.093 | 15.7 |
|  | 100 | 0.076 [0.056, 0.094] | -0.006 [-0.034, 0.017] | 0.031 | - |
|  | 1000 | 0.056 [0.036, 0.074] | -0.026 [-0.057, -0.001] | 0.003 | - |
| Land cover - urban areas (R) | 10 | 0.001 [0.000, 0.006] | -0.079 [-0.103, -0.063] | 0.000 | - |
|  | 100 | 0.001 [0.000, 0.005] | -0.081 [-0.104, -0.061] | 0.000 | - |
|  | 1000 | 0.002 [0.000, 0.006] | -0.081 [-0.103, -0.060] | 0.000 | - |
| Nightlights (2018, C) | 10 | 0.092 [0.068, 0.111] | 0.011 [-0.019, 0.032] | 0.077 | - |
|  | 100 | 0.049 [0.030, 0.070] | -0.033 [-0.058, -0.007] | 0.001 | - |
|  | 1000 | 0.012 [0.003, 0.028] | -0.069 [-0.091, -0.044] | 0.000 | - |
| Nightlights (2018, R) | 10 | 0.006 [0.002, 0.013] | -0.075 [-0.098, -0.057] | 0.000 | - |
|  | 100 | 0.001 [0.000, 0.005] | -0.080 [-0.106, -0.063] | 0.000 | - |
|  | 1000 | 0.000 [0.000, 0.004] | -0.081 [-0.106, -0.063] | 0.000 | - |
| Human population density (C) | **10** | **0.101 [0.080, 0.136]** | **0.021 [0.001, 0.043]** | **0.098** | **24.0** |
|  | 100 | 0.093 [0.072, 0.119] | 0.013 [-0.010, 0.030] | 0.090 | - |
|  | 1000 | 0.079 [0.058, 0.104] | -0.003 [-0.030, 0.021] | 0.041 | - |
| Human population density (R) | 10 | 0.011 [0.004, 0.018] | -0.070 [-0.093, -0.052] | 0.000 | - |
|  | 100 | 0.001 [0.000, 0.003] | -0.081 [-0.106, -0.062] | 0.000 | - |
|  | 1000 | 0.002 [0.000, 0.005] | -0.081 [-0.104, -0.061] | 0.000 | - |
| Expected number of primates of six | 10 | 0.068 [0.049, 0.106] | -0.012 [-0.032, 0.018] | 0.025 | - |
| genera (C) | 100 | 0.047 [0.031, 0.097] | -0.031 [-0.055, 0.012] | 0.016 | - |
|  | 1000 | 0.032 [0.012, 0.058] | -0.048 [-0.074, -0.024] | 0.000 | - |
| Expected number of primates of six | 10 | 0.053 [0.038, 0.071] | -0.028 [-0.048, -0.013] | 0.001 | - |
| genera (R) | 100 | 0.033 [0.020, 0.049] | -0.049 [-0.071, -0.031] | 0.000 | - |
|  | 1000 | 0.030 [0.017, 0.046] | -0.052 [-0.075, -0.034] | 0.000 | - |
