## Supplementary Data (Data S1 - S19) for "Climate and land-use shape the spread of zoonotic yellow fever virus": Data S11.docx

Data S11. Impact of several environmental factors on the dispersal velocity of YFV lineages in Brazil, based on the non-human YFV genomic data set and with environmental distances computed with the Circuitscape path model. The results are based on 100 posterior trees obtained by spatially-explicit phylogeographic inference. “C” and “R” indicate if the considered environmental raster was considered as a conductance (“C”) or resistance factor (“R”), and *k* is the rescaling parameter used to transform the initial raster (see the text for further details). For regression coefficients and *Q* values we report both the median estimate and the 95% HPD interval. The Bayes factor (BF) supports are only reported when p(*Q* > 0) is at least 90%. Following Kass & Raftery (1995), we consider a BF value >20 as strong support for a significant correlation between the environmental distances and dispersal durations (in bold), and a BF value >3 and <20 as positive (underlined).

| Environmental factor | *k* | Regression coefficient | *Q* statistic | p(*Q* > 0) | BF |
| --- | --- | --- | --- | --- | --- |
| *Aedes aegypti* (ENM, C) | 10 | 0.065 [0.046, 0.091] | -0.035 [-0.075, -0.010] | 0.01 | - |
|  | 100 | 0.054 [0.035, 0.081] | -0.045 [-0.090, -0.017] | 0.02 | - |
|  | 1000 | 0.052 [0.033, 0.080] | -0.046 [-0.092, -0.017] | 0.02 | - |
| *Aedes aegypti* ( ENM, R) | 10 | 0.070 [0.046, 0.118] | -0.027 [-0.044, -0.015] | 0.01 | - |
|  | 100 | 0.065 [0.041, 0.107] | -0.034 [-0.054, -0.019] | 0.00 | - |
|  | 1000 | 0.064 [0.040, 0.106] | -0.035 [-0.055, -0.020] | 0.00 | - |
| *Aedes albopictus* (C) | 10 | 0.089 [0.066, 0.146] | -0.008 [-0.023, 0.002] | 0.08 | - |
|  | 100 | 0.089 [0.067, 0.141] | -0.009 [-0.027, 0.003] | 0.12 | - |
|  | 1000 | 0.089 [0.067, 0.141] | -0.009 [-0.027, 0.003] | 0.12 | - |
| *Aedes albopictus* (R) | 10 | 0.099 [0.071, 0.155] | 0.000 [-0.013, 0.009] | 0.53 | - |
|  | 100 | 0.098 [0.070, 0.151] | -0.001 [-0.016, 0.008] | 0.43 | - |
|  | 1000 | 0.097 [0.070, 0.151] | -0.001 [-0.016, 0.008] | 0.42 | - |
| Annual mean temperature (C) | 10 | 0.119 [0.093, 0.179] | 0.020 [-0.004, 0.034] | 0.93 | 15.7 |
|  | 100 | 0.119 [0.093, 0.179] | 0.020 [-0.003, 0.035] | 0.94 | 15.7 |
|  | 1000 | 0.119 [0.094, 0.179] | 0.020 [-0.004, 0.035] | 0.94 | 15.7 |
| Annual mean temperature (R) | 10 | 0.102 [0.072, 0.173] | 0.003 [-0.026, 0.014] | 0.68 | - |
|  | 100 | 0.101 [0.070, 0.171] | 0.001 [-0.028, 0.013] | 0.59 | - |
|  | 1000 | 0.101 [0.070, 0.171] | 0.001 [-0.028, 0.013] | 0.57 | - |
| Annual precipitation (C) | 10 | 0.099 [0.075, 0.168] | 0.001 [-0.022, 0.014] | 0.52 | - |
|  | 100 | 0.095 [0.071, 0.163] | -0.003 [-0.026, 0.011] | 0.35 | - |
|  | 1000 | 0.094 [0.071, 0.162] | -0.004 [-0.026, 0.011] | 0.31 | - |
| Annual precipitation (R) | 10 | 0.113 [0.085, 0.170] | 0.015 [-0.014, 0.029] | 0.89 | - |
|  | 100 | 0.112 [0.085, 0.165] | 0.014 [-0.016, 0.029] | 0.86 | - |
|  | 1000 | 0.112 [0.085, 0.165] | 0.014 [-0.017, 0.029] | 0.86 | - |
| Aridity (C) | 10 | 0.086 [0.052, 0.142] | -0.017 [-0.039, 0.027] | 0.18 | - |
|  | 100 | 0.079 [0.047, 0.136] | -0.024 [-0.043, 0.022] | 0.17 | - |
|  | 1000 | 0.079 [0.047, 0.135] | -0.024 [-0.044, 0.021] | 0.17 | - |
| Aridity (R) | 10 | 0.104 [0.049, 0.146] | 0.005 [-0.039, 0.029] | 0.66 | - |
|  | 100 | 0.102 [0.045, 0.140] | 0.004 [-0.043, 0.025] | 0.59 | - |
|  | 1000 | 0.102 [0.044, 0.139] | 0.003 [-0.043, 0.024] | 0.58 | - |
| Distance to main roads (C) | 10 | 0.088 [0.054, 0.124] | -0.014 [-0.045, 0.013] | 0.13 | - |
|  | 100 | 0.025 [0.011, 0.041] | -0.075 [-0.132, -0.046] | 0.00 | - |
|  | 1000 | 0.001 [0.000, 0.005] | -0.098 [-0.164, -0.07] | 0.00 | - |
| Distance to main roads (R) | 10 | 0.105 [0.064, 0.161] | 0.002 [-0.028, 0.049] | 0.64 | - |
|  | 100 | 0.080 [0.050, 0.134] | -0.021 [-0.050, 0.018] | 0.09 | - |
|  | 1000 | 0.064 [0.037, 0.109] | -0.036 [-0.070, -0.003] | 0.02 | - |
| Elevation (C) | 10 | 0.034 [0.015, 0.061] | -0.066 [-0.108, -0.036] | 0.00 | - |
|  | 100 | 0.007 [0.001, 0.016] | -0.092 [-0.155, -0.062] | 0.00 | - |
|  | 1000 | 0.004 [0.000, 0.011] | -0.096 [-0.161, -0.064] | 0.00 | - |
| Elevation (R) | **10** | **0.124 [0.096, 0.164]** | **0.027 [-0.012, 0.055]** | **0.92** | **49.0** |
|  | 100 | 0.112 [0.084, 0.151] | 0.014 [-0.030, 0.047] | 0.77 | - |
|  | 1000 | 0.109 [0.080, 0.147] | 0.011 [-0.034, 0.045] | 0.69 | - |
| Enhanced vegetation index | 10 | 0.071 [0.041, 0.116] | -0.028 [-0.055, -0.003] | 0.01 | - |
| (2005-2018, C) | 100 | 0.060 [0.034, 0.095] | -0.039 [-0.071, -0.017] | 0.00 | - |
|  | 1000 | 0.058 [0.033, 0.093] | -0.041 [-0.073, -0.019] | 0.00 | - |
| Enhanced vegetation index | 10 | 0.103 [0.071, 0.158] | 0.002 [-0.022, 0.050] | 0.62 | - |
| (2005-2018, R) | 100 | 0.102 [0.070, 0.155] | 0.001 [-0.024, 0.047] | 0.57 | - |
|  | 1000 | 0.102 [0.070, 0.155] | 0.001 [-0.024, 0.047] | 0.56 | - |
| Forest loss (2000-2019, C) | 10 | 0.087 [0.062, 0.123] | -0.011 [-0.044, 0.014] | 0.21 | - |
|  | 100 | 0.031 [0.016, 0.055] | -0.067 [-0.125, -0.043] | 0.00 | - |
|  | 1000 | 0.004 [0.000, 0.015] | -0.095 [-0.161, -0.067] | 0.00 | - |
| Forest loss (2000-2019, R) | 10 | 0.090 [0.060, 0.134] | -0.009 [-0.041, 0.027] | 0.19 | - |
|  | 100 | 0.039 [0.023, 0.067] | -0.060 [-0.102, -0.027] | 0.00 | - |
|  | 1000 | 0.028 [0.014, 0.050] | -0.070 [-0.118, -0.039] | 0.00 | - |
| Land cover - herbaceous (C) | 10 | 0.019 [0.010, 0.031] | -0.077 [-0.138, -0.054] | 0.00 | - |
|  | 100 | 0.001 [0.000, 0.007] | -0.097 [-0.163, -0.070] | 0.00 | - |
|  | 1000 | 0.000 [0.000, 0.003] | -0.099 [-0.166, -0.070] | 0.00 | - |
| Land cover - herbaceous (R) | 10 | 0.060 [0.037, 0.096] | -0.037 [-0.086, -0.010] | 0.01 | - |
|  | 100 | 0.041 [0.021, 0.072] | -0.057 [-0.114, -0.030] | 0.00 | - |
|  | 1000 | 0.037 [0.018, 0.067] | -0.060 [-0.119, -0.033] | 0.00 | - |
| Land cover - mosaic | 10 | 0.059 [0.033, 0.089] | -0.040 [-0.084, -0.015] | 0.00 | - |
| natural vegetation (C) | 100 | 0.022 [0.008, 0.049] | -0.075 [-0.129, -0.045] | 0.00 | - |
|  | 1000 | 0.008 [0.001, 0.026] | -0.091 [-0.149, -0.060] | 0.00 | - |
| Land cover - mosaic | 10 | 0.044 [0.028, 0.067] | -0.054 [-0.104, -0.031] | 0.00 | - |
| natural vegetation (R) | 100 | 0.030 [0.017, 0.046] | -0.068 [-0.122, -0.043] | 0.00 | - |
|  | 1000 | 0.027 [0.015, 0.043] | -0.070 [-0.126, -0.045] | 0.00 | - |
| Land cover - urban areas (C) | 10 | 0.099 [0.073, 0.155] | 0.000 [-0.021, 0.025] | 0.52 | - |
|  | 100 | 0.076 [0.053, 0.116] | -0.019 [-0.055, -0.002] | 0.02 | - |
|  | 1000 | 0.052 [0.030, 0.080] | -0.045 [-0.091, -0.024] | 0.00 | - |
| Land cover - urban areas (R) | 10 | 0.001 [0.000, 0.007] | -0.098 [-0.164, -0.069] | 0.00 | - |
|  | 100 | 0.002 [0.000, 0.008] | -0.096 [-0.165, -0.067] | 0.00 | - |
|  | 1000 | 0.003 [0.000, 0.010] | -0.094 [-0.164, -0.065] | 0.00 | - |
| Nightlights (2018, C) | 10 | 0.093 [0.067, 0.126] | -0.007 [-0.054, 0.020] | 0.42 | - |
|  | 100 | 0.044 [0.021, 0.069] | -0.054 [-0.115, -0.019] | 0.00 | - |
|  | 1000 | 0.012 [0.003, 0.025] | -0.086 [-0.153, -0.056] | 0.00 | - |
| Nightlights (2018, R) | 10 | 0.009 [0.003, 0.020] | -0.091 [-0.154, -0.062] | 0.00 | - |
|  | 100 | 0.002 [0.000, 0.008] | -0.096 [-0.164, -0.069] | 0.00 | - |
|  | 1000 | 0.001 [0.000, 0.007] | -0.096 [-0.165, -0.07] | 0.00 | - |
| Human population density (C) | 10 | 0.108 [0.074, 0.166] | 0.008 [-0.016, 0.047] | 0.83 | - |
|  | 100 | 0.098 [0.074, 0.143] | 0.001 [-0.024, 0.033] | 0.55 | - |
|  | 1000 | 0.085 [0.058, 0.119] | -0.012 [-0.05, 0.015] | 0.18 | - |
| Human population density (R) | 10 | 0.010 [0.003, 0.021] | -0.089 [-0.151, -0.063] | 0.00 | - |
|  | 100 | 0.001 [0.000, 0.007] | -0.097 [-0.165, -0.066] | 0.00 | - |
|  | 1000 | 0.003 [0.000, 0.010] | -0.095 [-0.164, -0.064] | 0.00 | - |
| Expected number of primates of six | 10 | 0.074 [0.041, 0.145] | -0.026 [-0.052, 0.029] | 0.17 | - |
| genera (C) | 100 | 0.022 [0.007, 0.073] | -0.075 [-0.104, -0.033] | 0.00 | - |
|  | 1000 | 0.003 [0.000, 0.018] | -0.094 [-0.154, -0.067] | 0.00 | - |
| Expected number of primates of six | 10 | 0.075 [0.055, 0.100] | -0.024 [-0.065, 0.004] | 0.05 | - |
| genera (R) | 100 | 0.050 [0.034, 0.069] | -0.047 [-0.101, -0.018] | 0.00 | - |
|  | 1000 | 0.046 [0.031, 0.065] | -0.051 [-0.105, -0.021] | 0.00 | - |
